# Viral dynamics of SARS-CoV-2 variants in vaccinated and unvaccinated individuals

**DOI:** 10.1101/2021.02.16.21251535

**Authors:** Stephen M. Kissler, Joseph R. Fauver, Christina Mack, Caroline G. Tai, Mallery I. Breban, Anne E. Watkins, Radhika M. Samant, Deverick J. Anderson, Jessica Metti, Gaurav Khullar, Rachel Baits, Matthew MacKay, Daisy Salgado, Tim Baker, Joel T. Dudley, Christopher E. Mason, David D. Ho, Nathan D. Grubaugh, Yonatan H. Grad

## Abstract

**Background:** The alpha and delta SARS-CoV-2 variants have been responsible for major recent waves of COVID-19 despite increasing vaccination rates. The reasons for the increased transmissibility of these variants and for the reduced transmissibility of vaccine breakthrough infections are unclear.

**Methods:** We quantified the course of viral proliferation and clearance for 173 individuals with acute SARS-CoV-2 infections using longitudinal quantitative RT-PCR tests conducted using anterior nares/oropharyngeal samples (*n =* 199,941) as part of the National Basketball Association’s (NBA) occupational health program between November 28^th^, 2020, and August 11^th^, 2021. We measured the duration of viral proliferation and clearance and the peak viral concentration separately for individuals infected with alpha, delta, and non-variants of interest/variants of concern (non-VOI/VOC), and for vaccinated and unvaccinated individuals.

**Results:** The mean viral trajectories of alpha and delta infections resembled those of non-VOI/VOC infections. Vaccine breakthrough infections exhibited similar proliferation dynamics as infections in unvaccinated individuals (mean peak Ct: 20.5, 95% credible interval [19.0, 21.0] *vs*. 20.7 [19.8, 20.2], and mean proliferation time 3.2 days [2.5, 4.0] *vs*. 3.5 days [3.0, 4.0]); however, vaccinated individuals exhibited faster clearance (mean clearance time: 5.5 days [4.6, 6.6] *vs*. 7.5 days [6.8, 8.2]).

**Conclusions:** Alpha, delta, and non-VOI/VOC infections feature similar viral trajectories. Acute infections in vaccinated and unvaccinated people feature similar proliferation and peak Ct, but vaccinated individuals cleared the infection more quickly. Viral concentrations do not fully explain the differences in infectiousness between SARS-CoV-2 variants, and mitigation measures are needed to limit transmission from vaccinated individuals.

---

Two opposing forces shaping the COVID-19 pandemic are (1) the emergence of increasingly transmissible SARS-CoV-2 variants of concern (VOCs) and (2) the uptake of vaccines that prevent infection, protect against severe disease, and reduce transmission. Among the VOCs, of special interest are the alpha (lineage B.1.1.7) and delta (B.1.617.2, AY.1, AY.2, AY.3, and AY.3.1) variants, responsible for recent waves of COVID-19.^1^ These variants feature mutations in the spike protein receptor binding domain^2^ that may enhance ACE-2 binding,^3^ thus increasing the efficiency of virus transmission. In addition to, and perhaps due to, these attributes, the viral trajectories for infections with alpha and delta could feature a higher peak viral load or longer duration of carriage, both of which could increase transmissibility. Meanwhile, preliminary evidence suggests that individuals with vaccine breakthrough infections are less likely to transmit,^4,5^ but whether this is attributable to lower peak viral loads, shorter duration of carriage, or both, remains uncertain.

By measuring viral concentration over the course of acute infection, it is possible to inform hypotheses about the mechanisms that underlie variation in transmissibility. Recent evidence suggests that delta-variant infections may feature substantially higher peak viral concentrations relative to other lineages,^6^ while viral concentrations in alpha-variant infections were indistinguishable from non-variant infections.^7^ Vaccinated individuals who become infected with SARS-CoV-2 may clear their infections more quickly than unvaccinated individuals,^8^ and vaccine breakthrough infections with delta may feature similar peak viral concentrations as non-breakthrough delta infections.^9^ However, many of these studies rely on cross-sectional viral concentration measurements triggered by the onset of symptoms, which miss viral dynamics during the critical early stages of infection. Furthermore, population transmission dynamics can bias cross-sectional viral concentration measurements,^10^ making it difficult to compare viral concentrations between variants that emerged at different periods of the pandemic.

To overcome these limitations, we collected and analyzed a prospective, longitudinal set of SARS-CoV-2 viral samples from 173 individuals obtained as part of the National Basketball Association’s occupational health program. Using a Bayesian hierarchical statistical model, we compared SARS-CoV-2 viral dynamics between individuals infected with alpha, delta, and non-variants of interest/variants of concern (non-VOI/VOCs) as well as for vaccinated and unvaccinated individuals.

## Methods

### Study design

The data reported here represent a convenience sample including team staff, players, arena staff, and other vendors (e.g., transportation, facilities maintenance, and food preparation) affiliated with the National Basketball Association (NBA). The study period ran between November 28^th^, 2020, and August 11^th^, 2021. Clinical samples were obtained by combined swabs of the anterior nares and oropharynx administered by a trained provider. Viral concentration was measured using the cycle threshold (Ct) according to the Roche cobas target 1 assay. Ct values were converted to viral genome equivalents using a standard curve (**Supplementary methods**).

### Study oversight

In accordance with the guidelines of the Yale Human Investigations Committee, this work with de-identified samples was approved for research not involving human subjects by the Yale Institutional Review Board (HIC protocol # 2000028599). This project was designated exempt by the Harvard Institutional Review Board (IRB20-1407).

### Study participants

Out of an initial pool of 872 participants who tested positive for SARS-CoV-2 infection during the study period, 173 individuals (90% male) had clinically confirmed novel infections that met our inclusion criteria: at least three positive PCR tests (Ct < 40), at least one negative PCR test (Ct = 40), and at least one test with Ct < 32 with the first positive test (Ct <40) occurring before August 1^st^ to ensure full sampling of the trajectory before the end of the study period. (**Table 1**). A total of 19,941 samples were available for this cohort, averaging 548 samples per week. Of the individuals who met the inclusion criteria, 36 were infected with alpha (B.1.1.7) and 36 with delta (B.1.617.2, AY.1, AY.2, AY.3, or AY.3.1), as confirmed by sequencing. An additional 28 individuals were infected with other variants of interest/variants of concern. There were 37 individuals with vaccine breakthrough infections, defined as infections for which the first positive test occurred at least two weeks after receipt of the final dose. Of these, 23 received the Pfizer-BioNTech vaccine, 8 received the Johnson & Johnson/Janssen vaccine, and 3 received the Moderna vaccine. The vaccine manufacturer was not reported for the remaining 3 individuals.

**Table 1.**
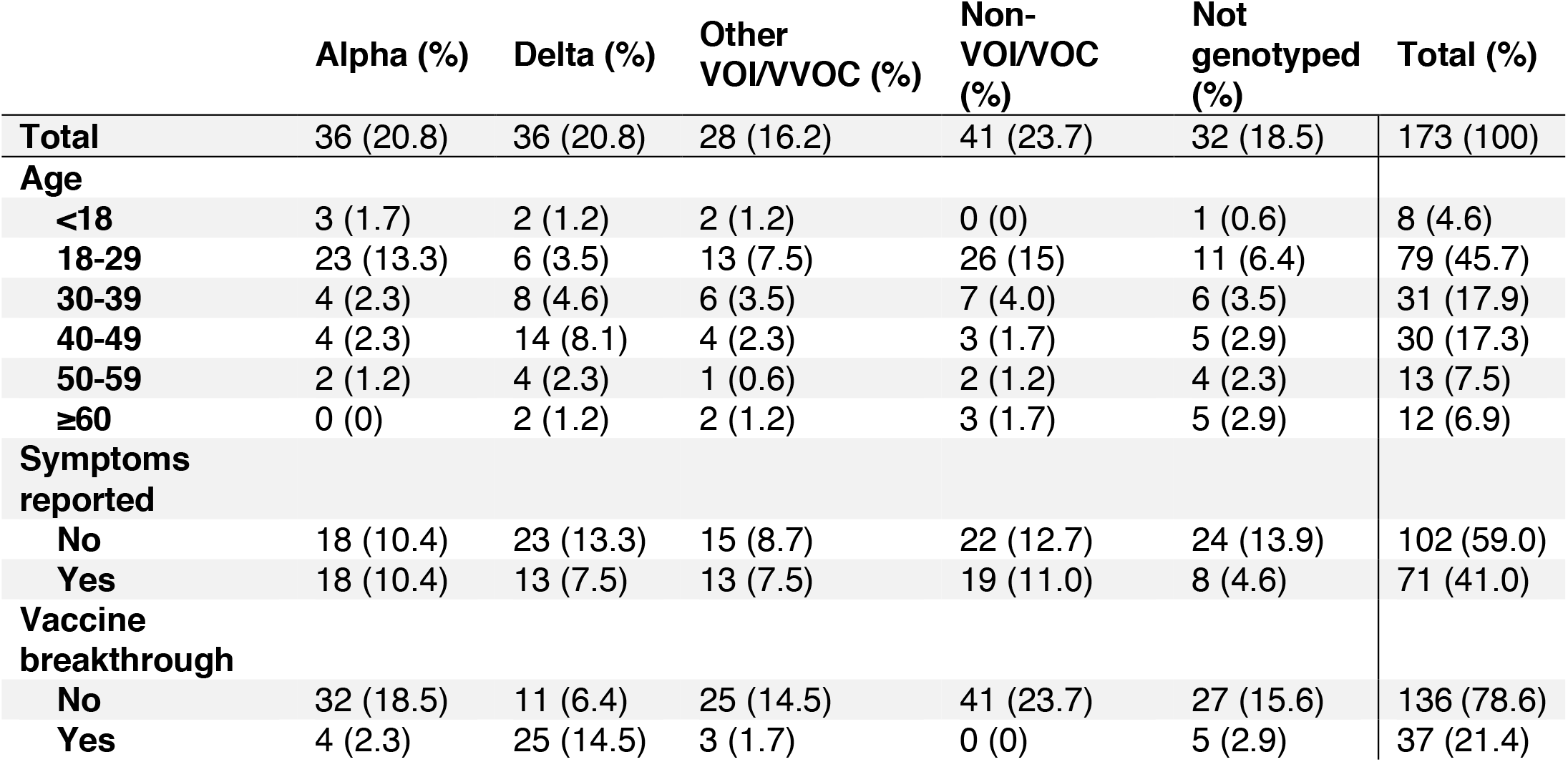
Characteristics of the study population. Number and percent (in parentheses) of individuals in the study population by age group, reported symptoms, and vaccine breakthrough status, stratified by variant.

### Study outcomes

We quantified the viral proliferation duration (time from first possible detection to peak viral concentration) the viral clearance duration (time from peak viral concentration to clearance of acute infection), the duration of acute infection (proliferation duration plus clearance duration), and the peak viral concentration for each person. We also quantified the population mean values of these quantities separately for individuals infected with alpha (*n* = 36), delta (*n* = 36), and non-VOI/VOCs (*n* = 41), as well as for vaccinated (*n* = 37) and unvaccinated (*n* = 136) individuals.

### Genome sequencing and lineage assignments

RNA was extracted and confirmed as SARS-CoV-2 positive by RT-qPCR with the Thermo Fisher TaqPath SARS-CoV-2 assay.^11^ Next Generation Sequencing with the Illumina COVIDSeq ARTIC primer set^12^ was used for viral amplification. Library preparation was performed using the amplicon-based Illumina COVIDseq Test v03^13^ and sequenced 2×74 on Illumina NextSeq 550 following the protocol as described in Illumina’s documentation.^14^ The resulting FASTQs were processed and analyzed on Illumina BaseSpace Labs using the Illumina DRAGEN COVID Lineage Application;^15^ versions included are 3.5.0, 3.5.1, 3.5.2, and 3.5.3. The DRAGEN COVID Lineage pipeline was run with default parameters recommended by Illumina. Samples were considered SARS-COV-2 positive if at least 5 viral amplicon targets were detected at 20x coverage. Each SARS-COV-2 positive sample underwent lineage assignment and phylogenetics analysis using the most updated version of Pangolin^16^ and NextClade,^17^ respectively.

### Statistical analysis

Following previously described methods,^18^ we used a Bayesian hierarchical model to estimate the proliferation duration, clearance duration, and peak viral concentration for each person and for the sub-populations of interest. The model describes the log_10_ viral concentration during an acute infection using a continuous piecewise-linear curve with control points that specify the time of acute infection onset, the time and magnitude of peak viral concentration, and the time of acute infection clearance. The assumption of piecewise linearity is equivalent to assuming exponential viral growth during the proliferation period followed by exponential viral decay during the clearance period. The control points were inferred using the Hamiltonian Monte Carlo algorithm as implemented in Stan (version 2.24).^19^ We used priors informed by a previous analysis^18^ for the main analysis and conducted a sensitivity analysis using vague priors as well as a strongly biased set of priors to assess robustness to the choice of prior. Full details are given in the **Supplementary methods**. Data and code are available online.^20^

## Results

### Summary of viral concentration measurements and model fit

A median of 6 samples (IQR: [4, 9]) with Ct values that surpassed the limit of detection (Ct = 40) were recorded for each person. The raw viral concentration measurements are depicted in **Figure 1** for individuals infected with alpha, delta, and non-VOI/VOCs as well as for unvaccinated and vaccinated infected individuals. Many of the tail samples depicted in **Figure 1** reflect samples with high Ct value/low viral concentration after the conclusion of acute infection. As these were not the main object of study in this analysis, any tests that occurred after the conclusion of an individual’s acute infection (as specified by the statistical model) are depicted in lighter shades. Visually, the trajectories appear similar across variants and vaccination statuses. While there are fewer low-level positives following acute infection for those with vaccine breakthrough and delta infections, this may reflect the fact that delta and breakthrough infections were more likely to occur near the end of the study period, which may have led to censoring of these points, as well as the substantial overlap in these categories (see **Table 1**). The individual-level model fits are depicted **Supplementary Figures 1-9**. The Gelman R-hat statistic^21^ was less than 1.1 for all parameters, indicating good convergence. There were no divergent iterations, indicating good exploration of the parameter space.

**Figure 1.**
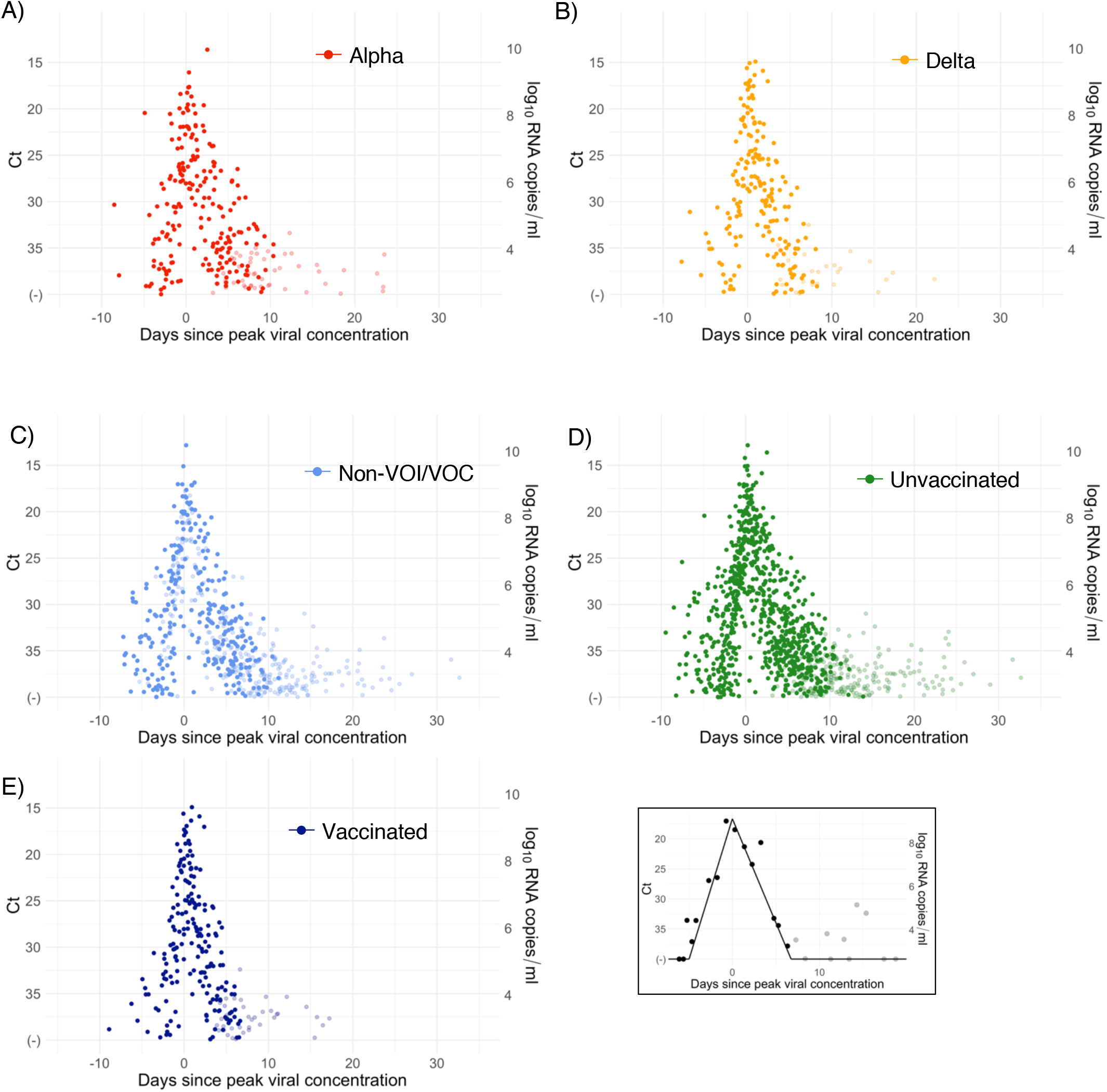
Raw Ct values by variant and vaccination status. Raw Ct values (points) for individuals infected with (A) alpha, (B) delta, or (C) non-VOI-VOCs, and for (D) unvaccinated and (E) vaccinated individuals. Points are horizontally aligned so that the inferred mean peak viral concentration for each person occurs at time 0. Points that fall after the conclusion of an individual’s acute infection, as measured by the individual’s mean posterior infection clearance time, are partially transparent, as these were not the focus of our study. The inset illustrates the process of making the tail points transparent: black points depict viral concentration measurements for a single person and the solid black lines depict the individual’s mean posterior viral trajectory.

### Viral trajectories by variant

We found no difference in the mean peak viral concentration, proliferation duration, clearance duration, or duration of acute infection for alpha or delta relative to non-VOI/VOCs, as evidenced by overlapping 95% credible intervals (**Figure 2A-F, Supplementary Table 1**). However, delta infections featured more frequent low peak Ct values, and corresponding high peak viral concentrations, than alpha or non-VOI/VOC infections, with 13.0% of the posterior delta trajectories surpassing a Ct value of 15 (9.6 log_10_ RNA copies/ml) *vs*. 6.9% and 10.2% of the posterior alpha and non-VOI/VOC trajectories surpassing the same threshold (**Figure 2G**). For those infected with delta, there is some evidence that vaccinated individuals tended to clear the virus more quickly than unvaccinated individuals (mean 5.9 days (95% credible interval [4.8, 7.2]) in vaccinated individuals *vs*. 7.6 days [5.5, 10.1] in unvaccinated individuals; **Supplementary Figure 10**), though the sample sizes are small and the 95% credible intervals for the mean clearance duration overlap.

**Figure 2.**
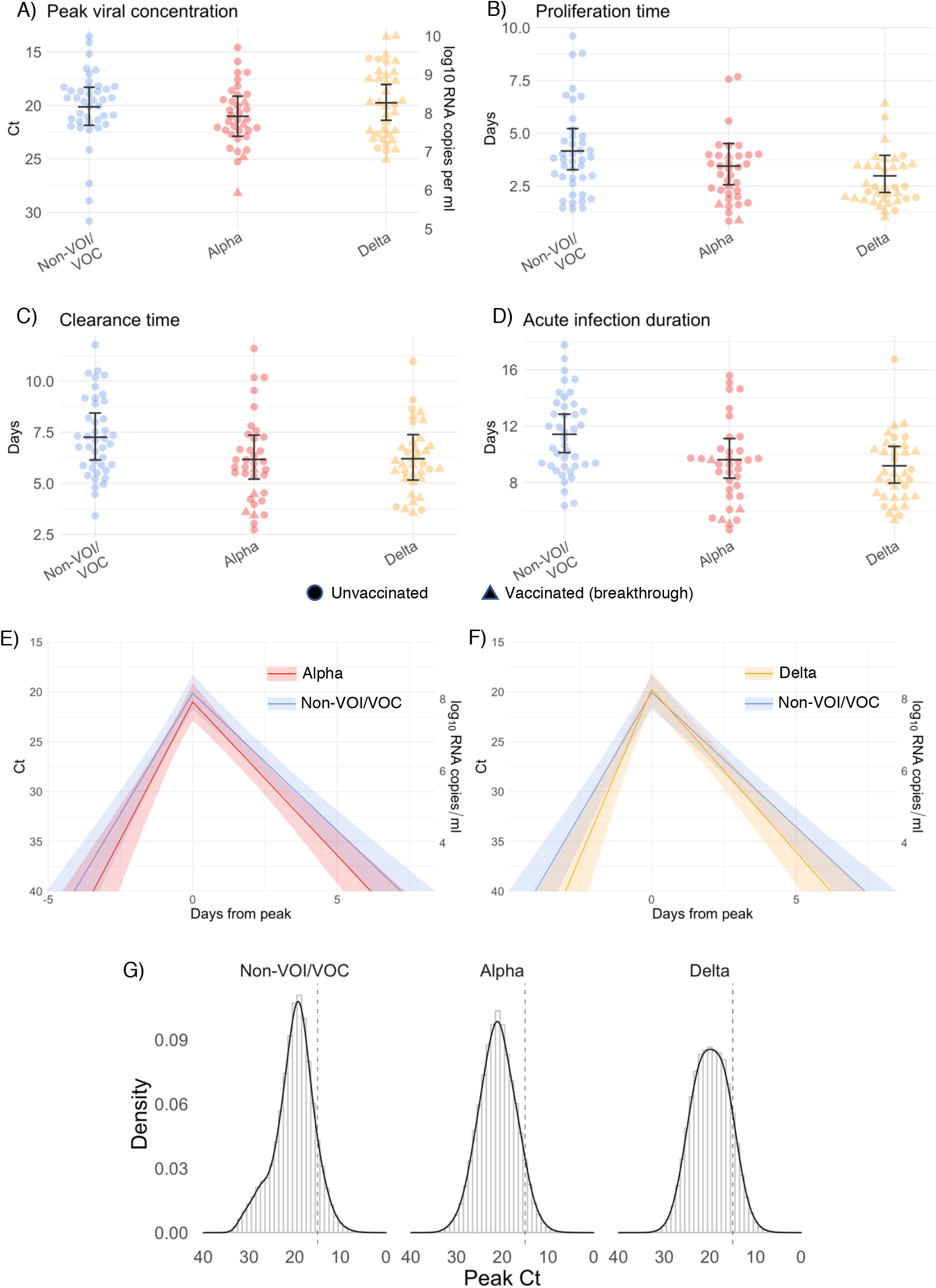
Estimated viral trajectory parameters for SARS-CoV-2 variants alpha and delta. Individual posterior means (points) with population means and 95% credible intervals (hatched lines) for (A) the peak viral concentration, (B) the proliferation duration, (C) the clearance duration, and (D) the total duration of acute infection for individuals infected with a non-VOI/VOC (light blue), alpha (red), or delta (orange). Circles denote unvaccinated individuals and triangles denote vaccinated individuals (breakthroughs). The points are jittered horizontally to avoid overlap. Panes (E)-(F) depict the mean posterior viral trajectories for alpha (E, red) and delta (F, orange) infections relative to non-VOI/VOC infections (light blue), as specified by the population means and credible intervals in (A)-(D). Solid lines in panes (E)-(F) depict the mean posterior viral trajectories and shaded regions represent 95% credible areas for the mean posterior trajectories. Histograms in pane (G) depict the posterior distributions of peak Ct values aggregated across all individuals infected with a non-VOI/VOC, alpha, and delta. The dashed line marks Ct = 15 (9.6 log_10_ RNA copies/ml) to facilitate comparison of the frequency of low peak Ct values/high peak viral concentrations across variants.

### Viral trajectories by vaccination status

We found no difference in the mean peak viral concentration or proliferation duration between vaccinated and unvaccinated individuals as evidenced by overlapping 95% credible intervals (**Figure 3**). However, breakthrough infections featured a faster clearance time (mean 5.5 days [4.6, 6.5] *vs*. 7.5 days [6.8, 8.2] in unvaccinated individuals), leading to a shorter overall duration of infection (8.7 days [7.6, 9.9] in vaccinated individuals *vs*. 11.0 days [10.3, 11.8] in unvaccinated individuals). We found no difference in viral trajectories for infected individuals who received the Pfizer-BioNTech vaccine (*n* = 23) vs. the Johnson & Johnson/Janssen vaccine (*n* = 8; **Supplementary Figure 11**). We did not assess viral trajectories for breakthrough infections in individuals who received the Moderna vaccine due to the small sample size (*n* = 3).

**Figure 3.**
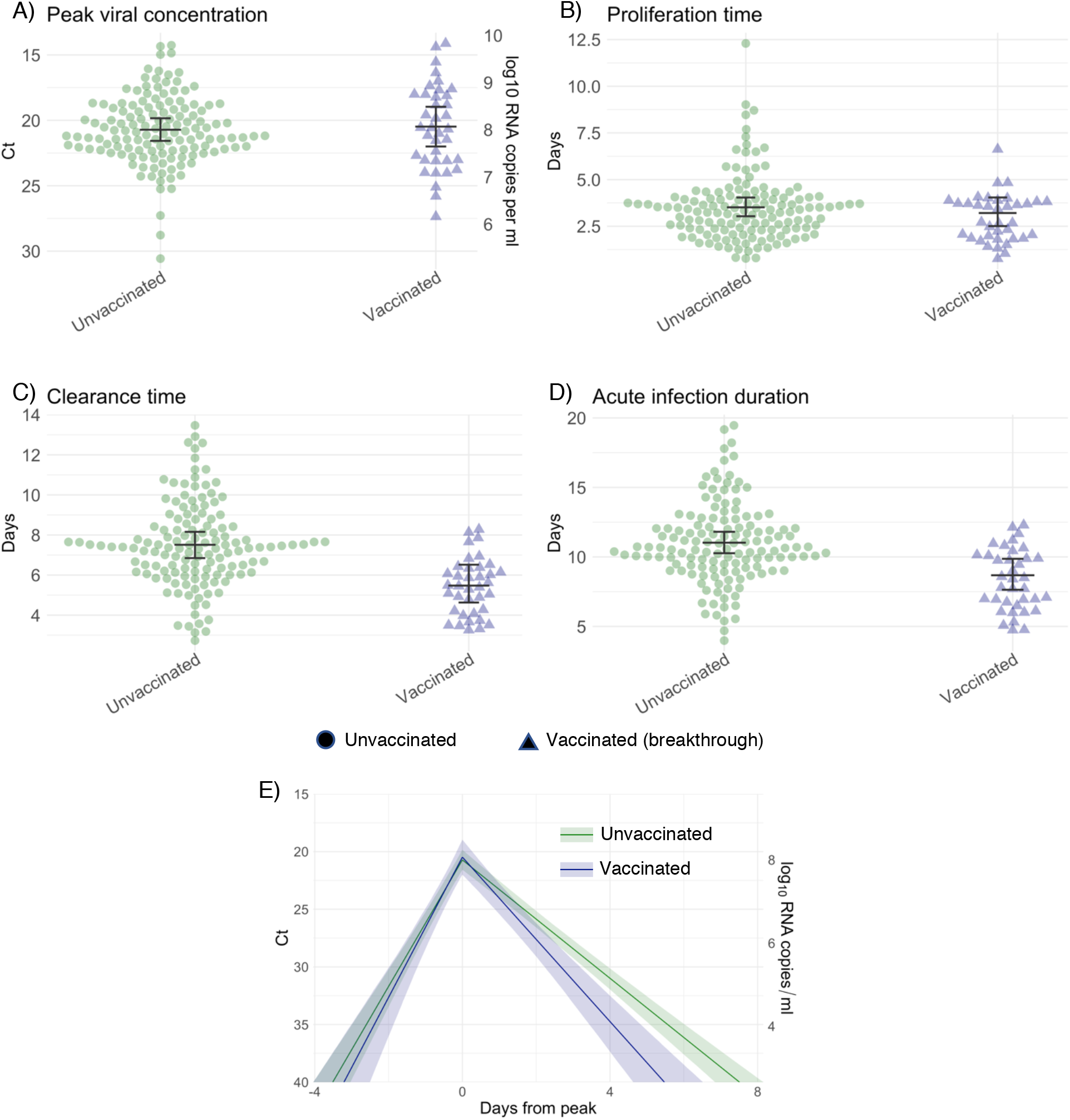
Estimated viral trajectory parameters for SARS-CoV-2 infections in unvaccinated and vaccinated individuals. Individual posterior means (points) with population means and 95% credible intervals (hatched lines) for (A) the peak viral concentration, (B) the proliferation duration, (C) the clearance duration, and (D) the total duration of acute infection for unvaccinated individuals (green) and vaccinated individuals (dark blue). Circles denote unvaccinated individuals and triangles denote vaccinated individuals (breakthroughs). The points are jittered horizontally to avoid overlap. Pane (E) depicts the mean posterior viral trajectories for vaccinated individuals (green) relative to unvaccinated individuals (dark blue), as specified by the population means and credible intervals in (A)-(D). Solid lines in pane (E) depict the mean posterior viral trajectories and shaded regions represent 95% credible areas for the mean posterior trajectories.

## Discussion

With the emergence of more transmissible SARS-CoV-2 variants such as alpha and delta, a key goal has been to understand which factors contribute to increased transmissibility. Our results indicate that the viral dynamics of infections caused by the alpha variant resemble those caused by the founding SARS-CoV-2 lineages, with similar proliferation and clearance times and similar peak viral concentrations. Viral dynamics in the oropharynx and nasopharynx therefore do not explain the elevated transmissibility of the alpha variant relative to the founding SARS-CoV-2 lineages.^22^ Instead, other factors, such as enhanced receptor binding which could lower the viral dose required for infection, may contribute to the alpha variant’s increased transmissibility. The viral dynamics of the delta variant are similar, with the exception that infections caused by the delta variant appear more likely to feature high peak viral concentrations. It is unclear if the greater proportion of cases with high peak viral concentrations reflects the underlying biology of the delta variant, the limited number of cases and sampling, or other factors, including the higher fraction of delta infections that were in vaccinated individuals. Infections with unusually high peak viral concentration may play an outsize role in spreading the virus, either by increasing the risk of transmission outside of close-contact settings^23^ or increasing the likelihood of “superspreading” events, pointing towards a possible mechanism for the enhanced transmissibility of the delta variant. Upper respiratory viral concentrations also do not explain the possible enhanced pathogenicity of the alpha and delta variants.^24^ Further studies are needed to uncover the origins of any differences in virulence, which could stem from differences in systemic viral dynamics that are not captured by oropharyngeal/nasopharyngeal samples.

A second key objective is to define the impact of COVID-19 vaccines on viral dynamics. Strong evidence demonstrates that each of the vaccines used by individuals in this cohort—the Pfizer/BioNTech, Moderna, and Johnson & Johnson/Janssen vaccines—reduces the rates of symptomatic COVID-19.^25–27^ A growing body of data also suggests that these vaccines reduce rates of asymptomatic infection.^28–30^ The extent to which infected vaccinated people can transmit SARS-CoV-2 has been unclear, with recent data supporting that breakthrough infections are infectious.^9^ Our data and a recent report from Singapore^8^ show that vaccine breakthrough cases follow a similar proliferation phase and reach similar peak viral concentrations as unvaccinated cases, but have a more rapid clearance phase, thereby modestly shortening the overall duration of infection. If the Ct values in vaccinated and unvaccinated infected individuals reflect the same amounts of infectious virus, then this implies that individuals with breakthrough infections may be as infectious as unvaccinated individuals in the early stage of the infection, but remain infectious for a shorter time, reducing the total degree of onward transmission. These findings are in keeping with the hypothesis that vaccination protects against the severe manifestations of disease but offers less protection against infection in the upper airway. Precautions are therefore necessary to prevent onward transmission even from vaccinated individuals.

Our ability to detect differences in SARS-CoV-2 viral dynamics between key populations was limited by small sample sizes and a high degree of interpersonal variation. More prospective longitudinal testing data within diverse cohorts is urgently needed to help resolve these patterns, particularly the peak viral concentration distribution for delta infections. The participants in this study were predominately young, male, and healthy, and therefore not representative across the general population. This underscores the need for similar studies in more diverse cohorts. Symptoms were not tracked throughout infection in this observational cohort; we were unable to assess differences in viral dynamics between symptomatic and asymptomatic individuals, nor were we able to link the timing of symptoms with key points in the viral trajectories. We did not test for the presence of infectious virus. While high viral concentrations are associated with elevated infectiousness,^31^ the nature of this association may be influenced by multiple factors, including variant, vaccination status, immune function, and host genetics.^32^ Viral culture studies and patient data would therefore help to contextualize the findings presented in this study.

This study provides a detailed picture of acute SARS-CoV-2 viral dynamics for key variants of concern in vaccinated and unvaccinated individuals. Frequent longitudinal measurements of viral concentrations can play a valuable role in illuminating factors contributing to SARS-CoV-2 transmissibility and the nature and extent of the impact of vaccination on viral dynamics in acute infections, thus informing interventions needed to mitigate the impact of COVID-19.

## Data Availability

Data and code are available online at https://github.com/gradlab/CtTrajectories_AllVariants

## Supplementary Appendix

### Converting Ct values to viral genome equivalents

To convert Ct values to viral genome equivalents, we first converted the Roche cobas target 1 Ct values to equivalent Ct values on a multiplexed version of the RT-qPCR assay from the US Centers for Disease Control and Prevention.^33^ We did this following our previously described methods.^18^ Briefly, we adjusted the Ct values using the best-fit linear regression between previously collected Roche cobas target 1 Ct values and CDC multiplex Ct values using the following regression equation:

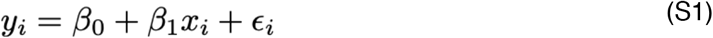

Here, *y*_*i*_ denotes the *i*^*th*^ Ct value from the CDC multiplex assay, *x*_*i*_ denotes the *i*^*th*^ Ct value from the Roche cobas target 1 test, and *ε*_*i*_ is an error term with mean 0 and constant variance across all samples. The coefficient values are *β*_*0*_ = –6.25 and *β*_*1*_ = 1.34.

Ct values were fitted to a standard curve to convert Ct value data to RNA copies. Synthetic T7 RNA transcripts corresponding to a 1,363 b.p. segment of the SARS-CoV-2 nucleocapsid gene were serially diluted from 10^6^-10^0^ RNA copies/μl in duplicate to generate a standard curve^34^ (**Supplementary Table 2**). The average Ct value for each dilution was used to calculate the slope (−3.60971) and intercept (40.93733) of the linear regression of Ct on log_10_ transformed standard RNA concentration, and Ct values from subsequent RT-qPCR runs were converted to RNA copies using the following equation:

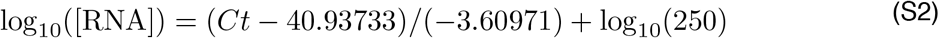

Here, [RNA] represents the RNA copies /ml. The log_10_(250) term accounts for the extraction (300 μl) and elution (75 μl) volumes associated with processing the clinical samples as well as the 1,000 μl/ml unit conversion.

### Model fitting

For the statistical analysis, we removed any sequences of 3 or more consecutive negative tests (Ct = 40) to avoid overfitting to these trivial values. Following our previously described methods,^18^ we assumed that the viral concentration trajectories consisted of a proliferation phase, with exponential growth in viral RNA concentration, followed by a clearance phase characterized by exponential decay in viral RNA concentration.^35^ Since Ct values are roughly proportional to the negative logarithm of viral concentration^36^, this corresponds to a linear decrease in Ct followed by a linear increase. We therefore constructed a piecewise-linear regression model to estimate the peak Ct value, the time from infection onset to peak (*i*.*e*. the duration of the proliferation stage), and the time from peak to infection resolution (*i*.*e*. the duration of the clearance stage). The trajectory may be represented by the equation

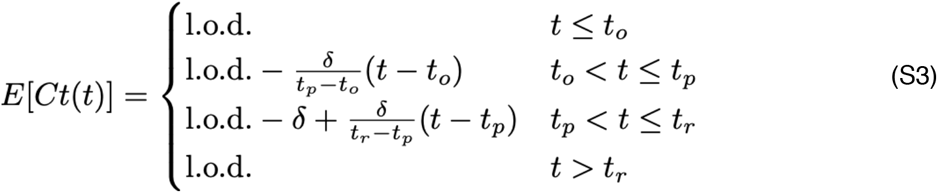

Here, E[*Ct(t)*] represents the expected value of the Ct at time *t*, “l.o.d” represents the RT-qPCR limit of detection, *δ* is the absolute difference in Ct between the limit of detection and the peak (lowest) Ct, and *t*_*o*_, *t*_*p*_, and *t*_*r*_ are the onset, peak, and recovery times, respectively.

Before fitting, we re-parametrized the model using the following definitions:

- Δ*Ct(t)* = l.o.d. – *Ct(t)* is the difference between the limit of detection and the observed Ct value at time *t*.
- *ω*_*p*_ *= t*_*p*_ *- t*_*o*_ is the duration of the proliferation stage.
- *ω*_*r*_ *= t*_*r*_ *- t*_*p*_ is the duration of the clearance stage.

We constrained 0.25 ≤ *ω*_*p*_ *≤* 14 days and 2 ≤ *ω*_*r*_ *≤* 30 days to prevent inferring unrealistically small or large values for these parameters for trajectories that were missing data prior to the peak and after the peak, respectively. We also constrained 0 ≤ *δ ≤* 40 as Ct values can only take values between 0 and the limit of detection (40).

We next assumed that the observed Δ*Ct(t)* could be described the following mixture model:

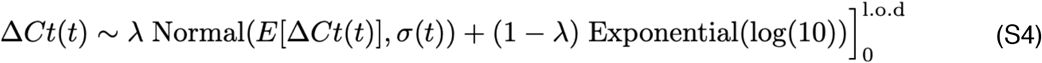

where E[Δ*Ct(t)*] = l.o.d. - E[*Ct(t)*] and *λ* is the sensitivity of the q-PCR test, which we fixed at 0.99. The bracket term on the right-hand side of the equation denotes that the distribution was truncated to ensure Ct values between 0 and the limit of detection. This model captures the scenario where most observed Ct values are normally distributed around the expected trajectory with standard deviation *σ(t)*, yet there is a small (1%) probability of an exponentially distributed false negative near the limit of detection. The log(10) rate of the exponential distribution was chosen so that 90% of the mass of the distribution sat below 1 Ct unit and 99% of the distribution sat below 2 Ct units, ensuring that the distribution captures values distributed at or near the limit of detection. We did not estimate values for *λ* or the exponential rate because they were not of interest in this study; we simply needed to include them to account for some small probability mass that persisted near the limit of detection to allow for the possibility of false negatives.

We used a hierarchical structure to describe the distributions of *ω*_*p*_, *ω*_*r*_, and *δ* for each person based on their respective population means *μ*_*ωp*_, *μ*_*ωr*_, and *μ*_*δ*_ and population standard deviations σ_ωp_, σ_ωr_, and σ_δ_ such that

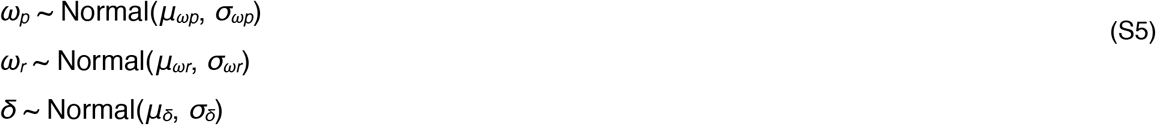

We inferred population means (*μ*_***•***_) separately for individuals infected with alpha, delta, and non-VOI/VOCs, as well as for unvaccinated and vaccinated individuals in a separate analysis. We used a Hamiltonian Monte Carlo fitting procedure implemented in Stan (version 2.24)^19^ and R (version 3.6.2)^37^ to estimate the individual-level parameters *ω*_*p*_, *ω*_*r*_, *δ*, and *t*_*p*_ as well as the population-level parameters *σ*, μ*_*ωp*_, *μ*_*ωr*_, *μ*_*δ*_, *σ*_*ωp*_, *σ*_*ωr*_, and *σ*_*δ*_. We used the following priors:

#### Hyperparameters

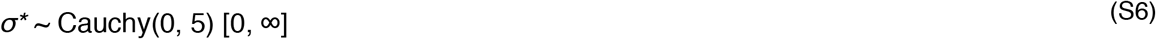

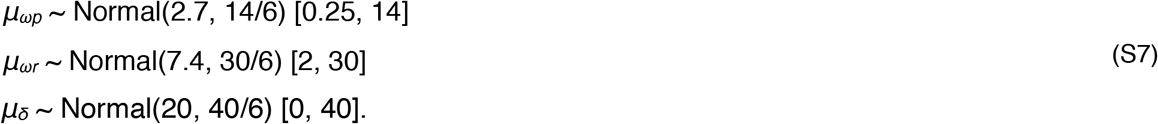

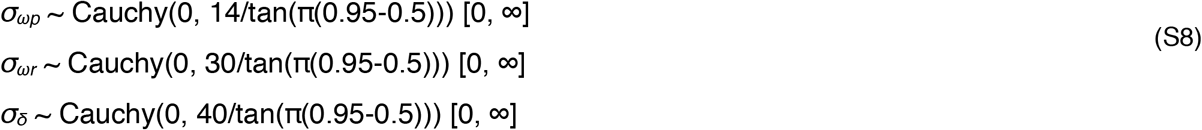

#### Individual-level parameters

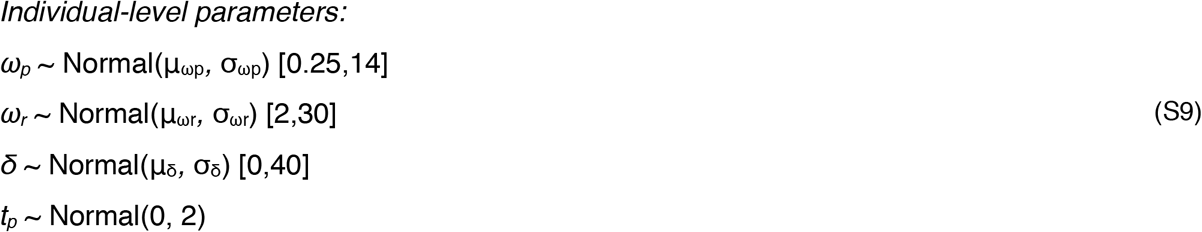

The values in square brackets denote truncation bounds for the distributions. We chose a vague half-Cauchy prior with scale 5 for the observation variance *σ\**. The priors for the population mean values (*μ*_***•***_) are normally distributed priors spanning the range of allowable values for that parameter; this prior is vague but expresses a mild preference for values near the posterior estimates obtained from a previous analysis.^38^ The priors for the population standard deviations (*σ*_***•***_) are half Cauchy-distributed with scale chosen so that 90% of the distribution sits below the maximum value for that parameter; this prior is vague but expresses a mild preference for standard deviations close to 0.

We ran four MCMC chains for 2,000 iterations each with a target average proposal acceptance probability of 0.8. The first half of each chain was discarded as the warm-up. The Gelman R-hat statistic was less than 1.1 for all parameters. This indicates good overall mixing of the chains. There were no divergent iterations, indicating good exploration of the parameter space. The posterior distributions for *μ*_*δ*_, *μ*_*ωp*_, and *μ*_*ωr*_, were estimated separately for individuals infected with alpha, delta, and non-VOI/VOCs as well as for vaccinated and unvaccinated individuals. These are depicted in **Figure 1** (main text). Draws from the individual posterior viral trajectory distributions are depicted in **Supplementary Figures 1-11**. The mean posterior viral trajectories for each person are depicted in **Supplementary Figure 12**.

### Assessing sensitivity to different priors

To ensure that our findings were not overly influenced by the prior distributions, we re-fit the model using two different sets of priors. The first “vague” set used posterior population means for *μ*_*ωp*_, *μ*_*ωr*_, and *μ*_*δ*_ chosen to lie near the center of the allowable range for those parameters. These priors were defined by

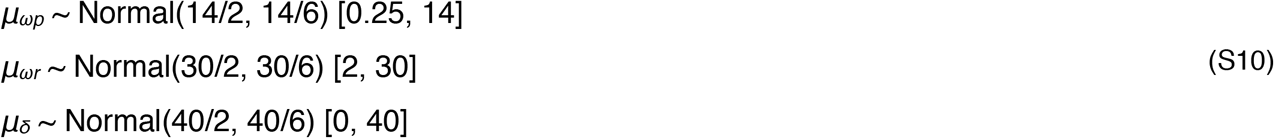

The second set used unrealistically low prior means for *μ*_*ωp*_, *μ*_*ωr*_, and *μ*_*δ*_ to check model robustness to highly biased prior distributions. These priors were defined by

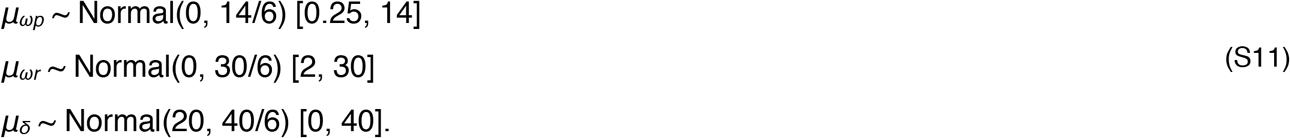

Note that we updated the prior means but kept the prior variances at their original wide values to avoid encoding over-confidence in the priors into the model. The posterior population means for these new sets of priors are depicted in **Supplementary Figures 13-14** (compare to **Figures 2-3**). Overall, the findings were consistent across choices of prior.

**Supplementary Table 1.**
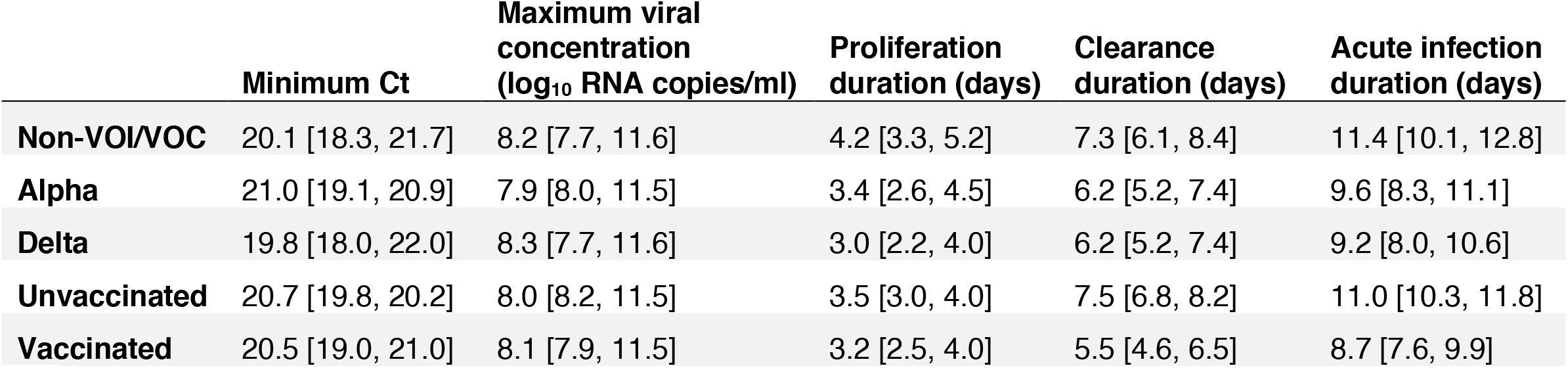
Posterior population viral trajectory parameters for SARS-CoV-2 infections by variant and vaccination status. Reported values represent the posterior mean and 95% credible intervals (brackets) for each parameter.

|  | Minimum Ct | Maximum viral<br>concentration<br>(log<sub>10</sub> RNA copies/ml) | Proliferation<br>duration (days) | Clearance<br>duration (days) | Acute infection<br>duration (days) |
| --- | --- | --- | --- | --- | --- |
| Non-VOI/VOC | 20.1 [18.3, 21.7] | 8.2 [7.7, 11.6] | 4.2 [3.3, 5.2] | 7.3 [6.1, 8.4] | 11.4 [10.1, 12.8] |
| Alpha | 21.0 [19.1, 20.9] | 7.9 [8.0, 11.5] | 3.4 [2.6, 4.5] | 6.2 [5.2, 7.4] | 9.6 [8.3, 11.1] |
| Delta | 19.8 [18.0, 22.0] | 8.3 [7.7, 11.6] | 3.0 [2.2, 4.0] | 6.2 [5.2, 7.4] | 9.2 [8.0, 10.6] |
| Unvaccinated | 20.7 [19.8, 20.2] | 8.0 [8.2, 11.5] | 3.5 [3.0, 4.0] | 7.5 [6.8, 8.2] | 11.0 [10.3, 11.8] |
| Vaccinated | 20.5 [19.0, 21.0] | 8.1 [7.9, 11.5] | 3.2 [2.5, 4.0] | 5.5 [4.6, 6.5] | 8.7 [7.6, 9.9] |

**Supplementary Table 2.** Standard curve relationship between virus RNA copies and Ct values. Synthetic T7 RNA transcripts corresponding to a 1,363 base pair segment of the SARS-CoV-2 nucleocapsid gene were serially diluted from 10^6^-10^0^ and evaluated in duplicate with RT-qPCR. The best-fit linear regression of the average Ct on the log_10_-transformed standard values had slope -3.60971 and intercept 40.93733 (R^2^ = 0.99).

| Standard<br>(copies/ul) | Replicate 1 (Ct) | Replicate 2 (Ct) | Average Ct |
| --- | --- | --- | --- |
| 10<sup>6</sup> | 19.3 | 19.7 | 19.5 |
| 10<sup>5</sup> | 23.0 | 21.2 | 22.1 |
| 10<sup>4</sup> | 26.9 | 26.7 | 26.8 |
| 10<sup>3</sup> | 30.6 | 30.4 | 30.5 |
| 10<sup>2</sup> | 34.0 | 34.0 | 34.0 |
| 10<sup>1</sup> | 37.2 | 36.6 | 36.9 |
| 10<sup>0</sup> | N/A | 39.9 | 39.9 |

**Supplementary Figure 1.**
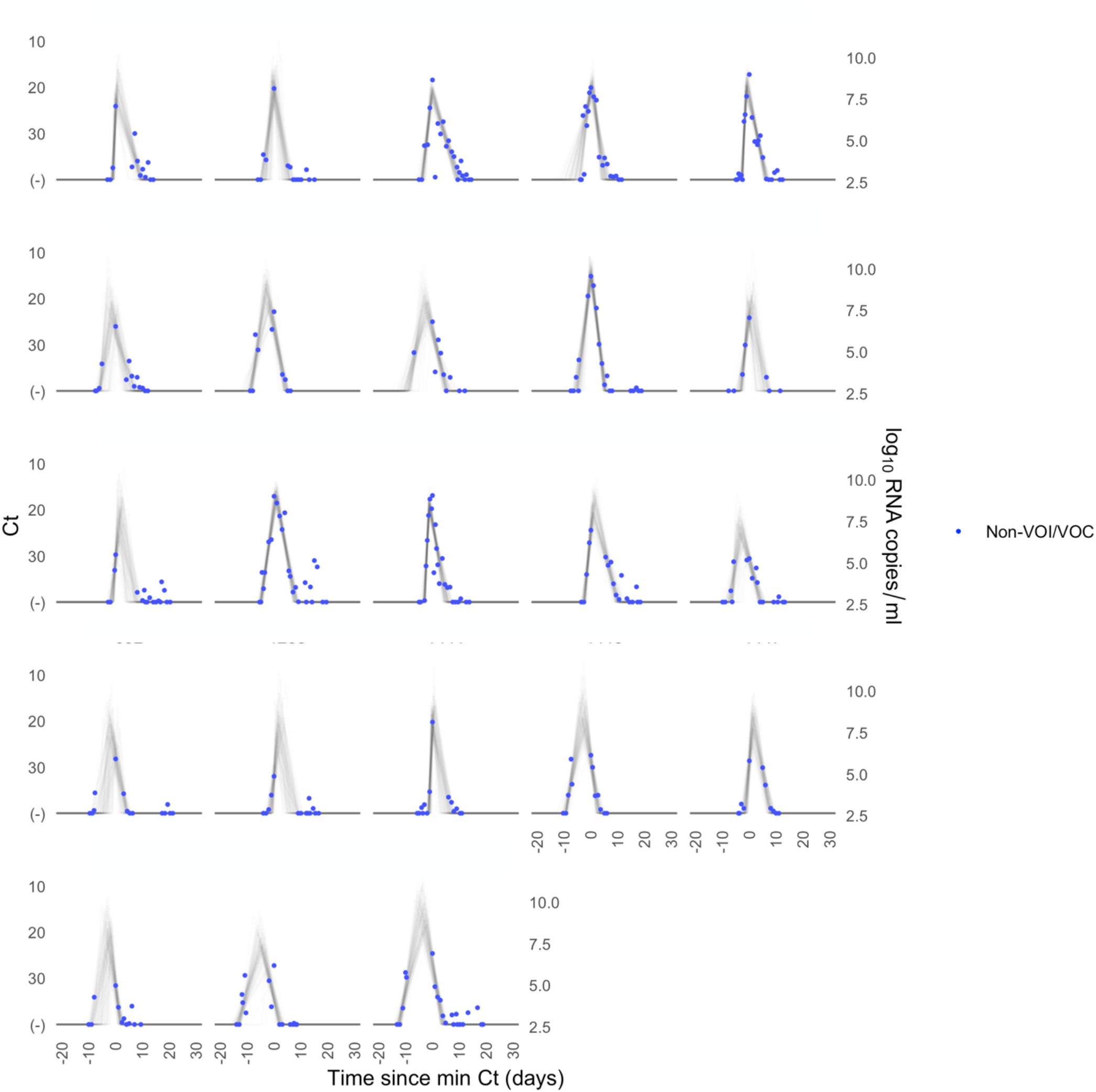
Ct values and estimated trajectories for non-VOI/VOC SARS-CoV-2 infections (1/3). Each pane depicts the recorded Ct values (points) and derived log-10 genome equivalents per ml (log(ge/ml)) for a single person during the study period. Points along the horizontal axis represent negative tests. Time is indexed in days since the minimum recorded Ct value (maximum viral concentration). Lines depict 100 draws from the posterior distribution for each person’s viral trajectory. Shaded boxes denote breakthrough infections.

**Supplementary Figure 2.**
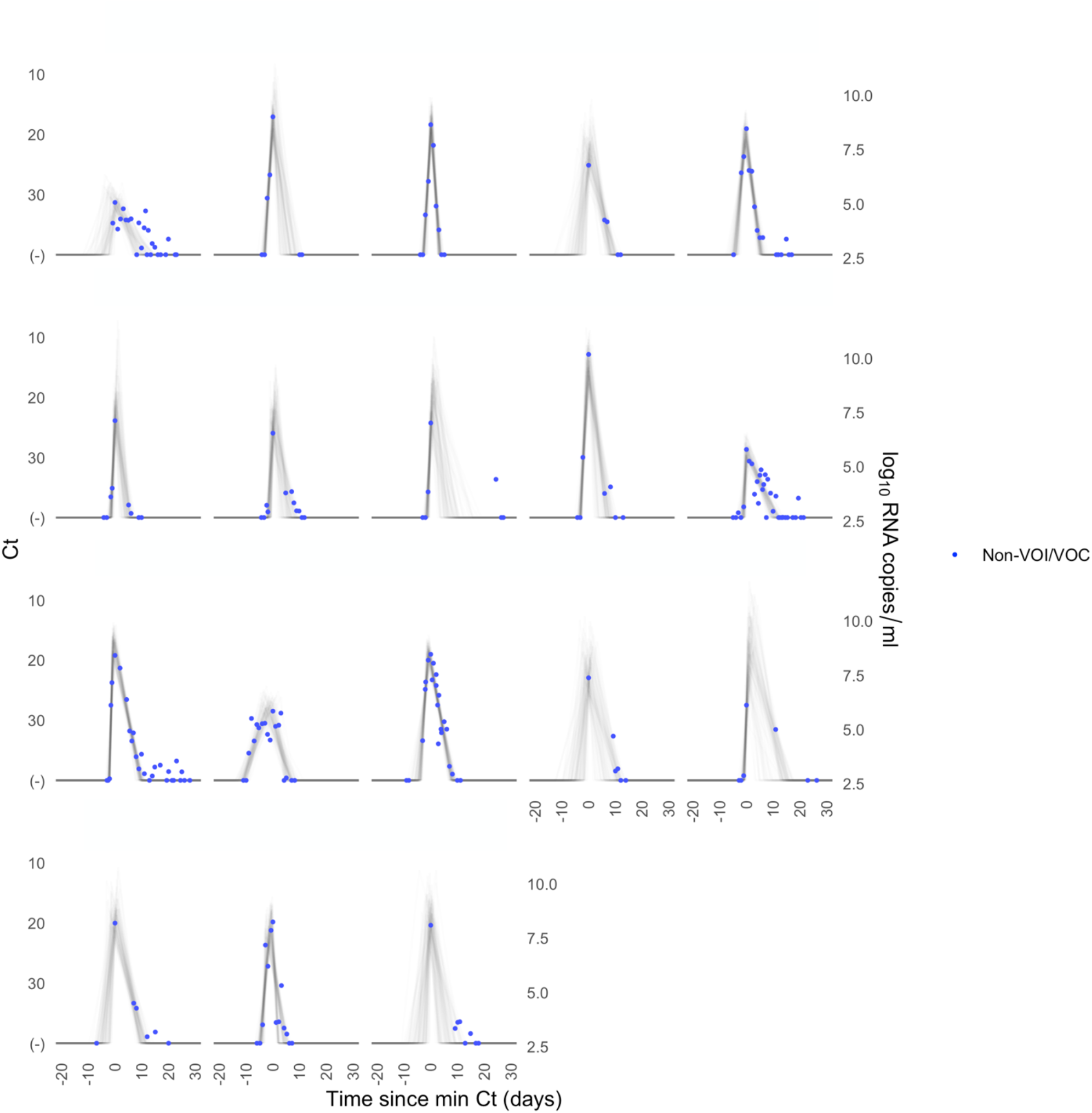
Ct values and estimated trajectories for non-VOI/VOC SARS-CoV-2 infections (2/3). Each pane depicts the recorded Ct values (points) and derived log-10 genome equivalents per ml (log(ge/ml)) for a single person during the study period. Points along the horizontal axis represent negative tests. Time is indexed in days since the minimum recorded Ct value (maximum viral concentration). Lines depict 100 draws from the posterior distribution for each person’s viral trajectory. Shaded boxes denote breakthrough infections.

**Supplementary Figure 3.**
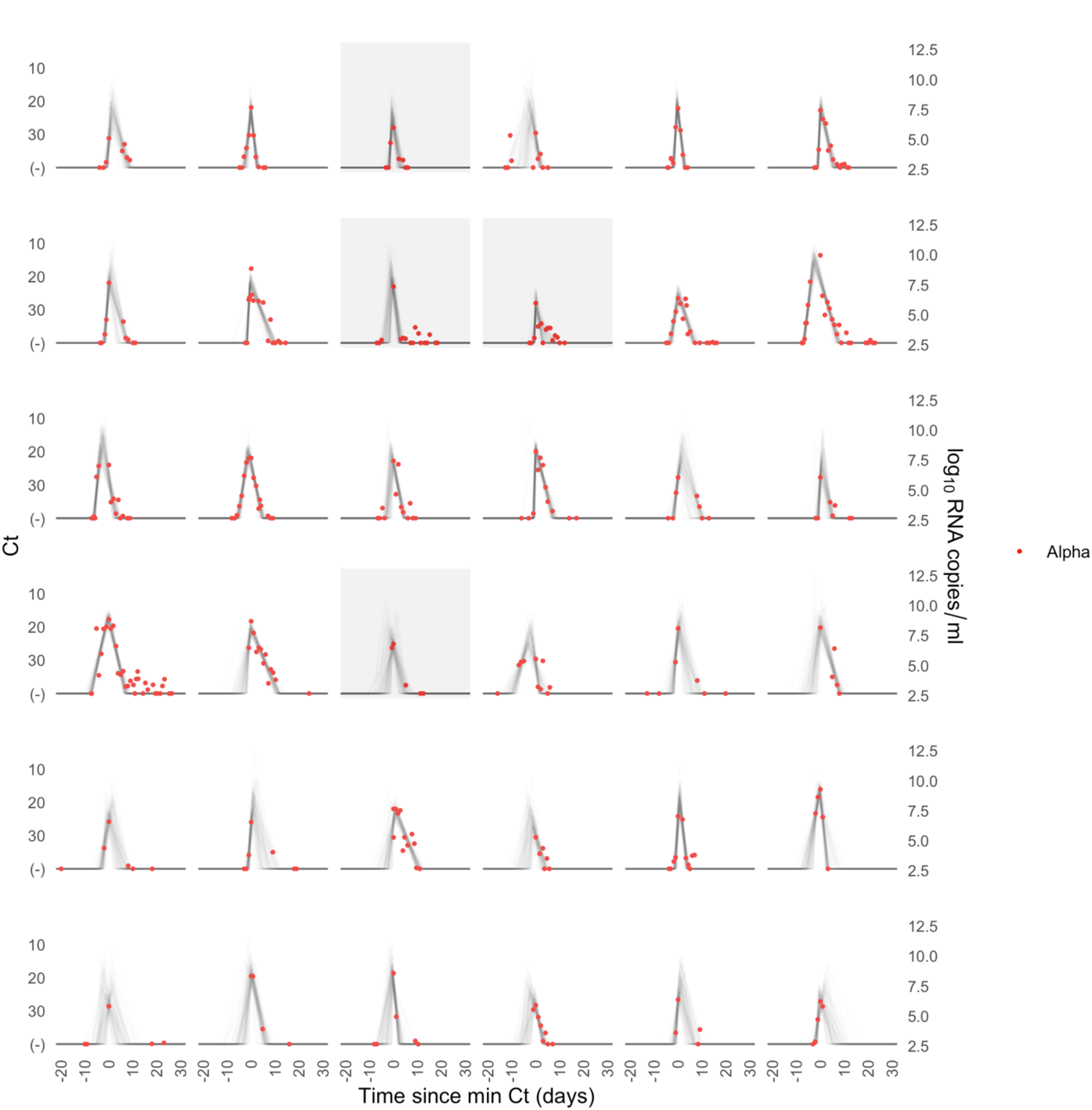
Ct values and estimated trajectories for alpha SARS-CoV-2 infections. Each pane depicts the recorded Ct values (points) and derived log-10 genome equivalents per ml (log(ge/ml)) for a single person during the study period. Points along the horizontal axis represent negative tests. Time is indexed in days since the minimum recorded Ct value (maximum viral concentration). Lines depict 100 draws from the posterior distribution for each person’s viral trajectory. Shaded boxes denote breakthrough infections.

**Supplementary Figure 4.**
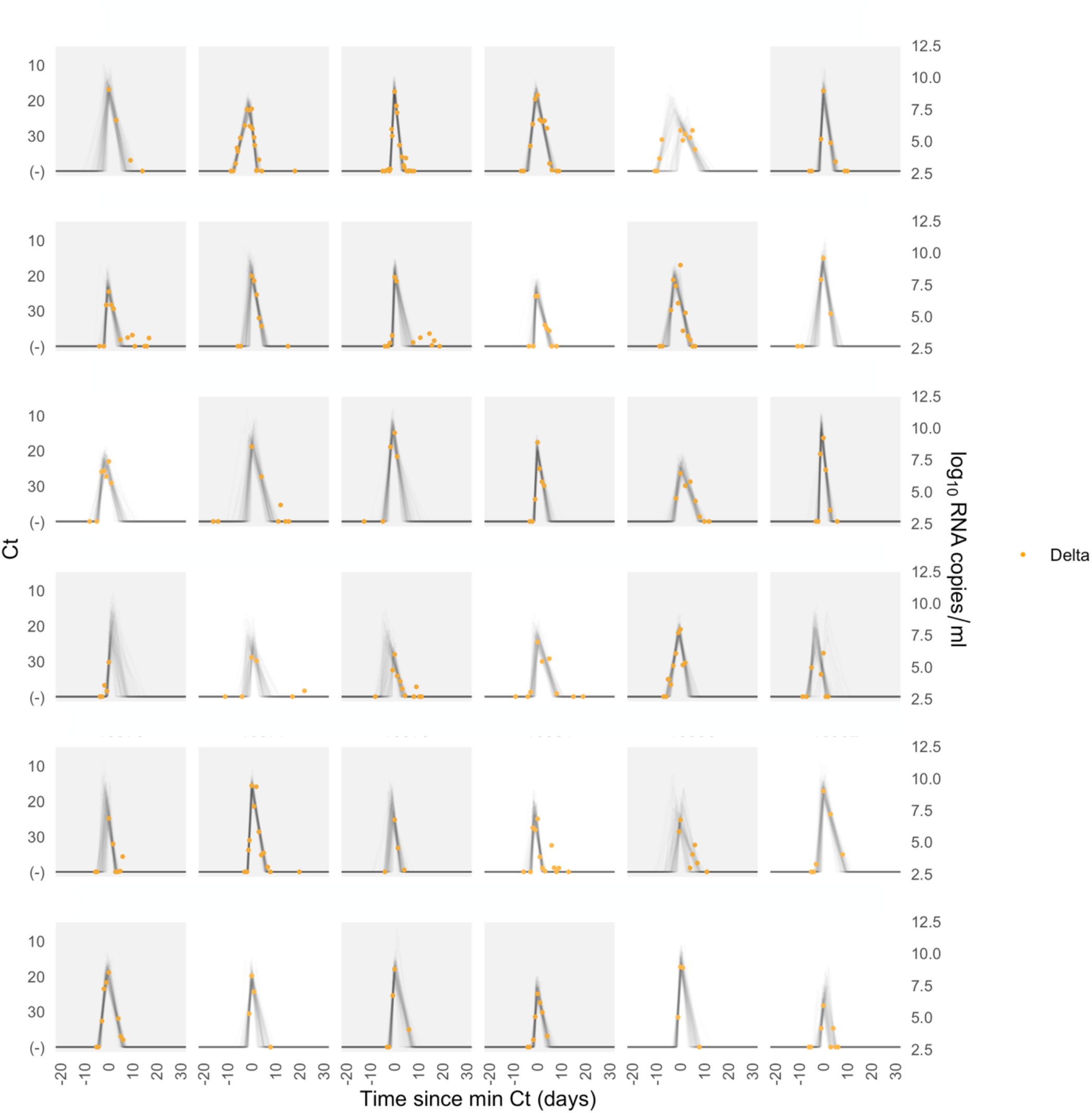
Ct values and estimated trajectories for delta SARS-CoV-2 infections. Each pane depicts the recorded Ct values (points) and derived log-10 genome equivalents per ml (log(ge/ml)) for a single person during the study period. Points along the horizontal axis represent negative tests. Time is indexed in days since the minimum recorded Ct value (maximum viral concentration). Lines depict 100 draws from the posterior distribution for each person’s viral trajectory. Shaded boxes denote breakthrough infections.

**Supplementary Figure 5.**
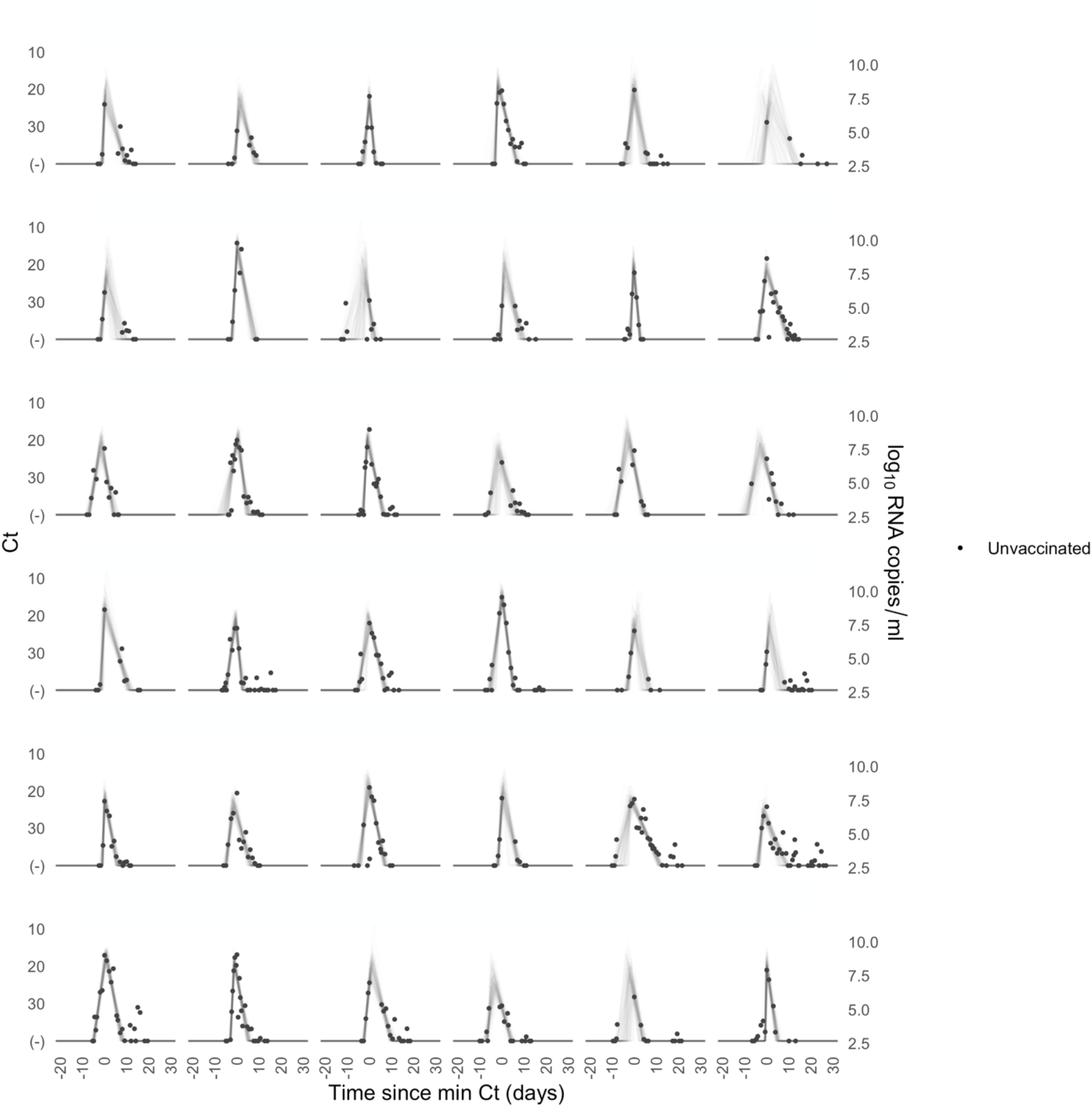
Ct values and estimated trajectories for SARS-CoV-2 infections in unvaccinated individuals (1/4). Each pane depicts the recorded Ct values (points) and derived log-10 genome equivalents per ml (log(ge/ml)) for a single person during the study period. Points along the horizontal axis represent negative tests. Time is indexed in days since the minimum recorded Ct value (maximum viral concentration). Lines depict 100 draws from the posterior distribution for each person’s viral trajectory.

**Supplementary Figure 6.**
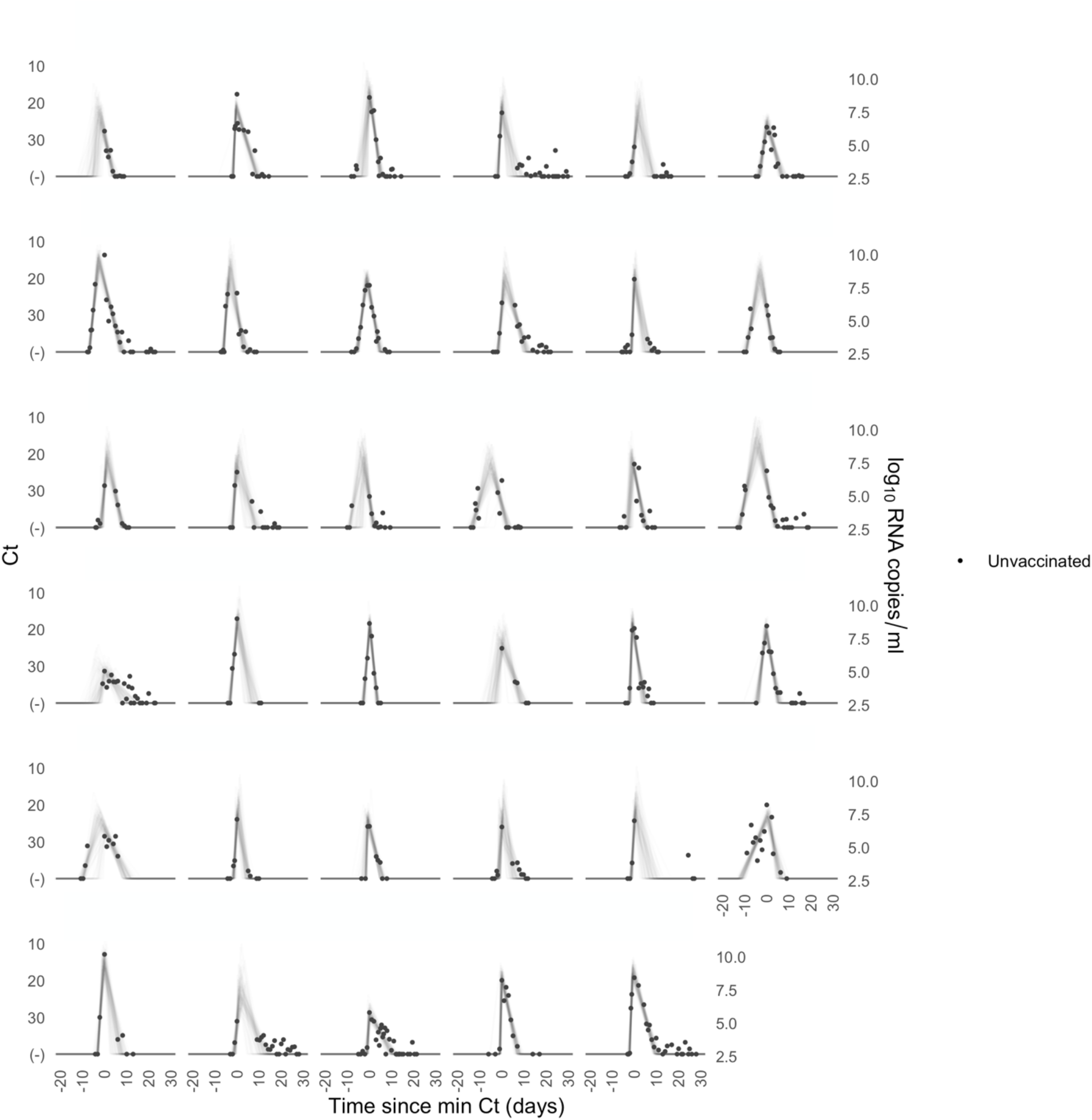
Ct values and estimated trajectories for SARS-CoV-2 infections in unvaccinated individuals (2/4). Each pane depicts the recorded Ct values (points) and derived log-10 genome equivalents per ml (log(ge/ml)) for a single person during the study period. Points along the horizontal axis represent negative tests. Time is indexed in days since the minimum recorded Ct value (maximum viral concentration). Lines depict 100 draws from the posterior distribution for each person’s viral trajectory.

**Supplementary Figure 7.**
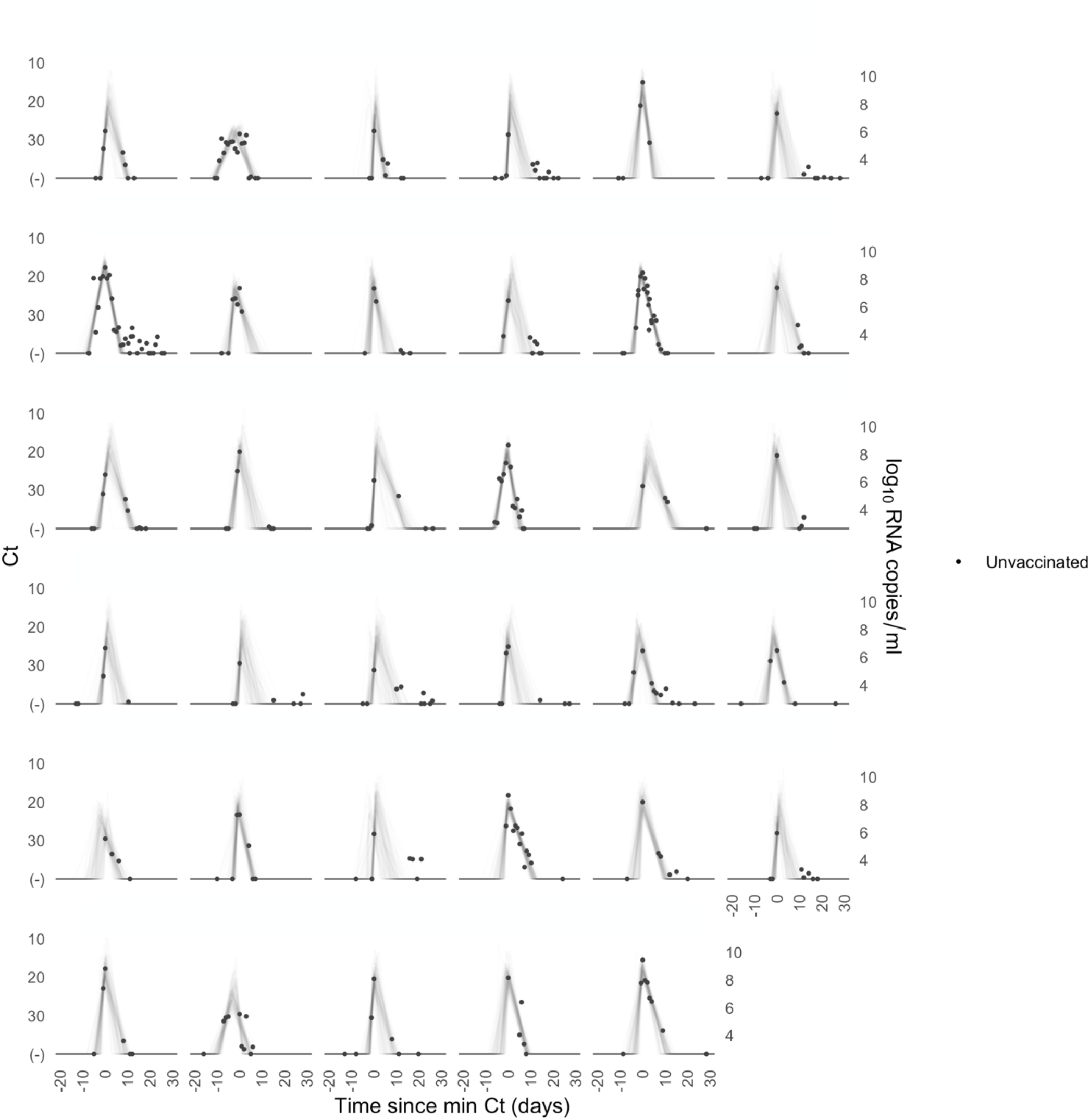
Ct values and estimated trajectories for SARS-CoV-2 infections in unvaccinated individuals (3/4). Each pane depicts the recorded Ct values (points) and derived log-10 genome equivalents per ml (log(ge/ml)) for a single person during the study period. Points along the horizontal axis represent negative tests. Time is indexed in days since the minimum recorded Ct value (maximum viral concentration). Lines depict 100 draws from the posterior distribution for each person’s viral trajectory.

**Supplementary Figure 8.**
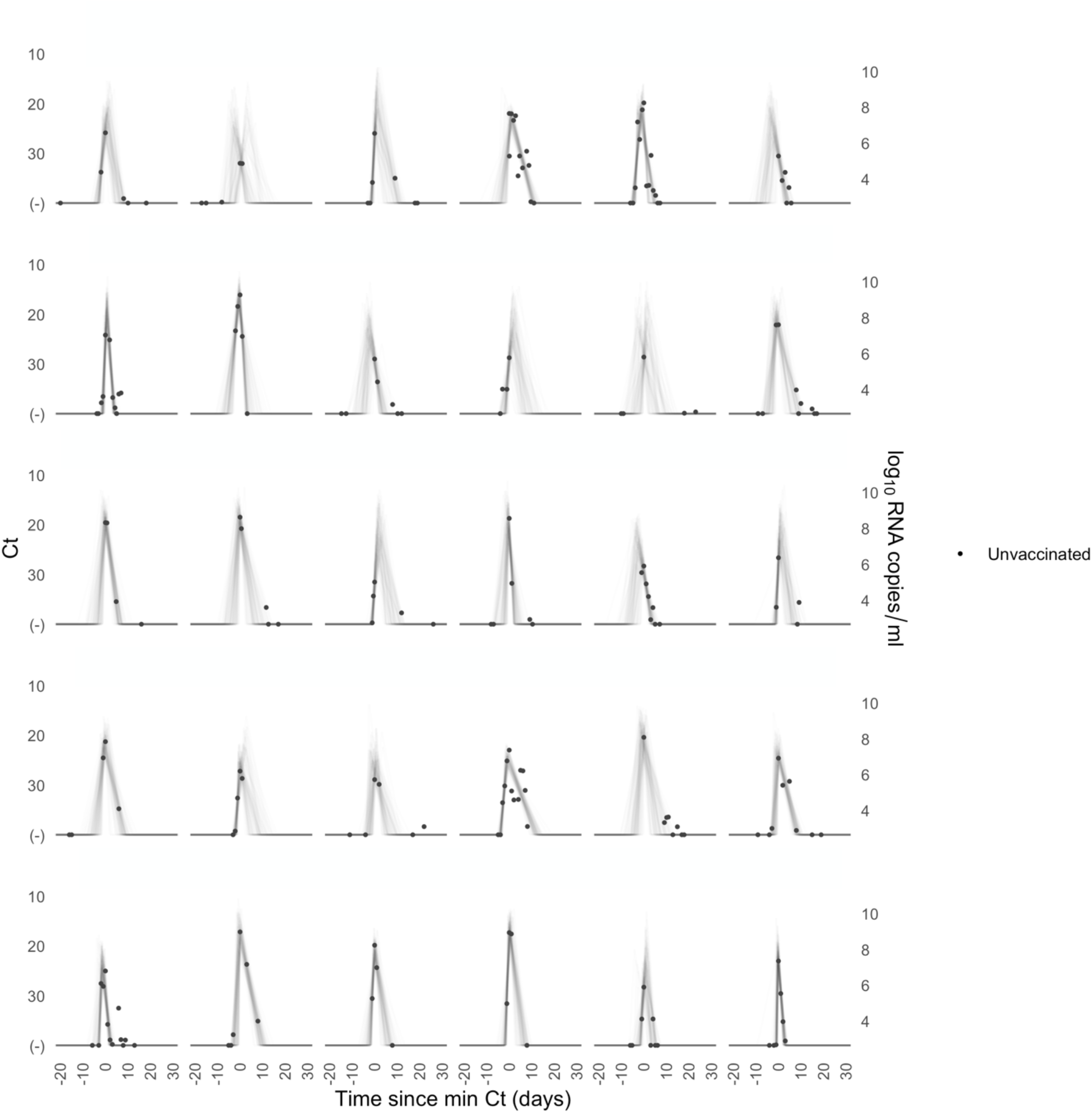
Ct values and estimated trajectories for SARS-CoV-2 infections in unvaccinated individuals (4/4). Each pane depicts the recorded Ct values (points) and derived log-10 genome equivalents per ml (log(ge/ml)) for a single person during the study period. Points along the horizontal axis represent negative tests. Time is indexed in days since the minimum recorded Ct value (maximum viral concentration). Lines depict 100 draws from the posterior distribution for each person’s viral trajectory.

**Supplementary Figure 9.**
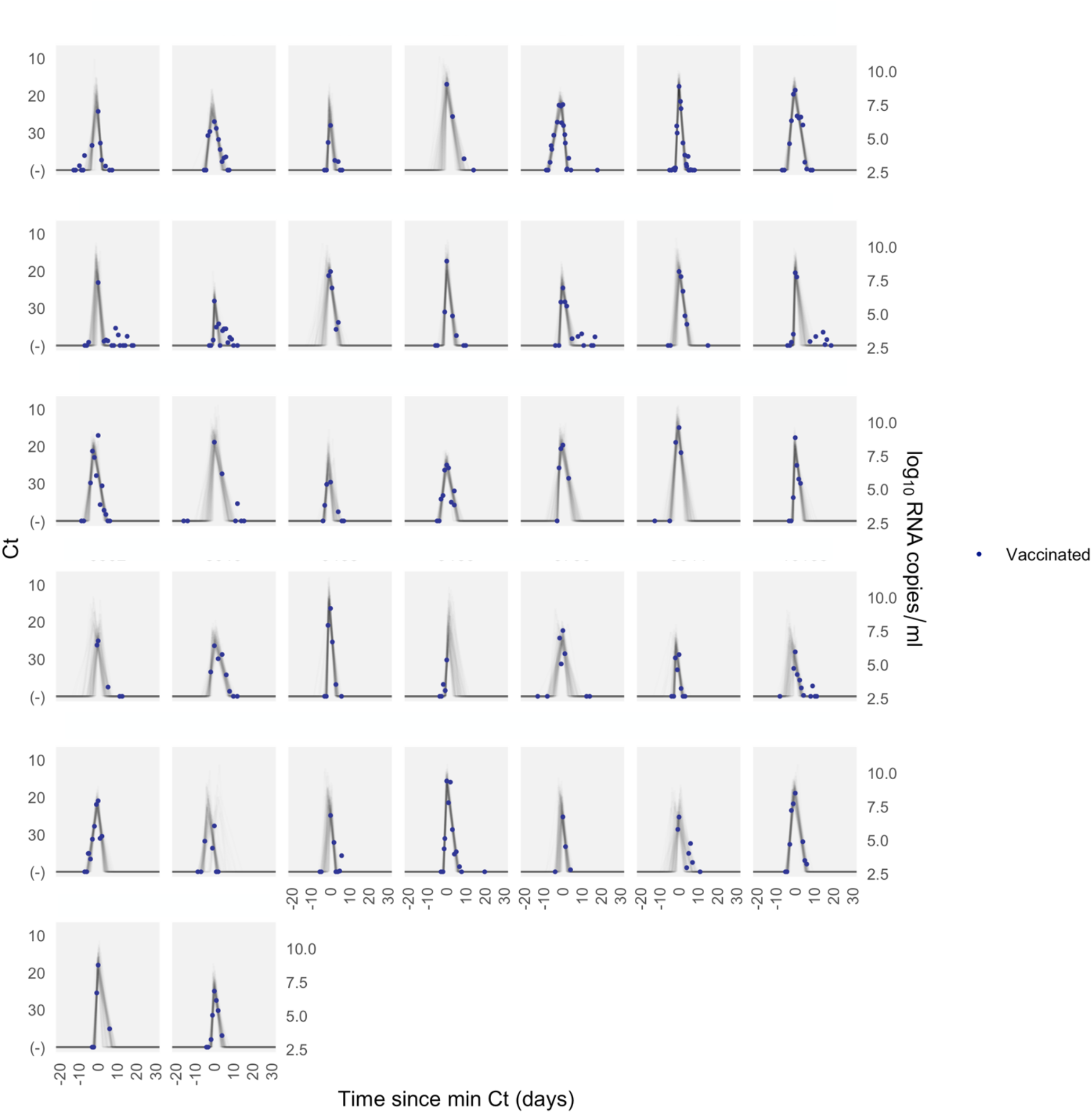
Ct values and estimated trajectories for SARS-CoV-2 infections in vaccinated individuals. Each pane depicts the recorded Ct values (points) and derived log-10 genome equivalents per ml (log(ge/ml)) for a single person during the study period. Points along the horizontal axis represent negative tests. Time is indexed in days since the minimum recorded Ct value (maximum viral concentration). Lines depict 100 draws from the posterior distribution for each person’s viral trajectory. Shaded boxes denote breakthrough infections.

**Supplementary Figure 10.**
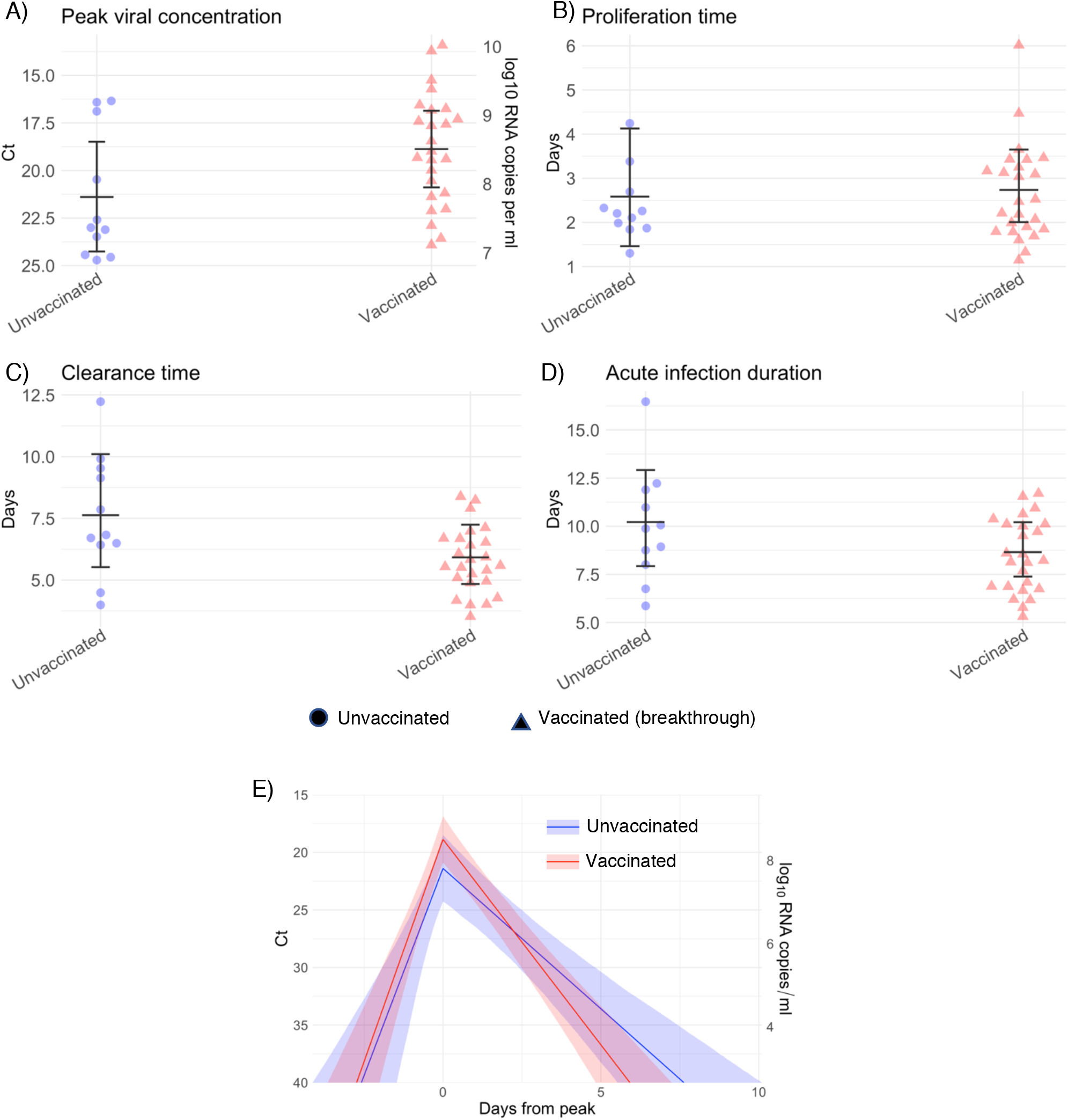
Estimated viral trajectory parameters for vaccinated and unvaccinated individuals infected with SARS-CoV-2 variant delta. Individual posterior means (points) with population means and 95% credible intervals (hatched lines) for (A) the peak viral concentration, (B) the proliferation duration, (C) the clearance duration, and (D) the total duration of acute infection for unvaccinated (blue) and vaccinated (red) individuals infected with delta. Circles denote unvaccinated individuals and triangles denote vaccinated individuals (breakthroughs). The points are jittered horizontally to avoid overlap. Pane (E) depicts the mean posterior viral trajectories for unvaccinated (blue) *vs*. vaccinated (red) individuals, as specified by the population means and credible intervals in (A)-(D). Solid lines in pane (E) depict the mean posterior viral trajectories and shaded regions represent 95% credible areas for the mean posterior trajectories.

**Supplementary Figure 11.**
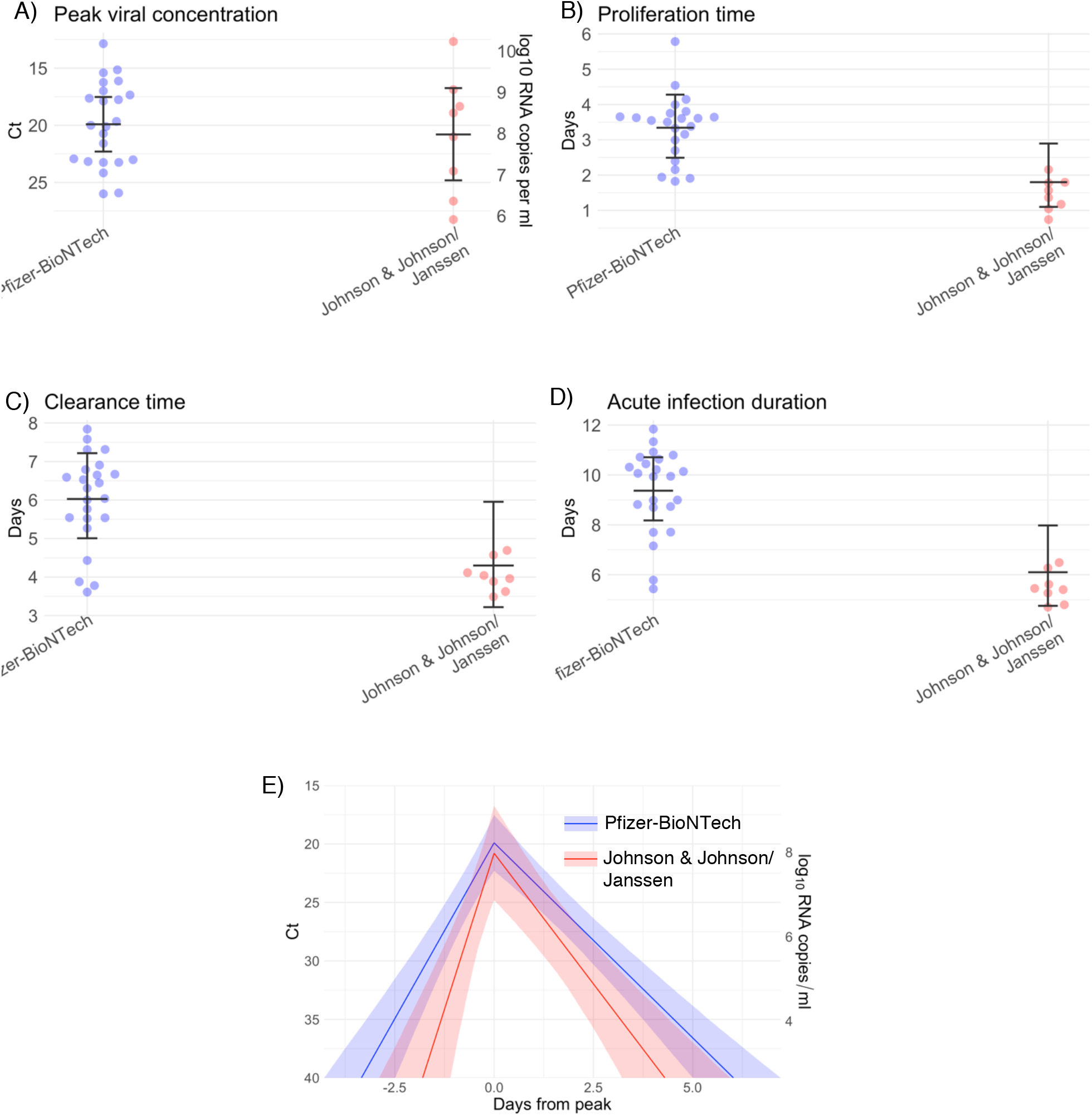
Estimated viral trajectory parameters for individuals vaccinated with the Pfizer-BioNTech vaccine vs. the Johnson & Johnson/Janssen vaccine. Individual posterior means (points) with population means and 95% credible intervals (hatched lines) for (A) the peak viral concentration, (B) the proliferation duration, (C) the clearance duration, and (D) the total duration of acute infection for breakthrough infections in individuals vaccinated with the Pfizer-BioNTech vaccine (blue) and the Johnson & Johnson/Janssen vaccine (red). The points are jittered horizontally to avoid overlap. Pane (E) depicts the mean posterior viral trajectories for breakthrough infections in individuals vaccinated with the Pfizer-BioNTech vaccine (blue) *vs*. the Johnson & Johnson/Janssen vaccine (red), as specified by the population means and credible intervals in (A)-(D). Solid lines in pane (E) depict the mean posterior viral trajectories and shaded regions represent 95% credible areas for the mean posterior trajectories.

**Supplementary Figure 12.**
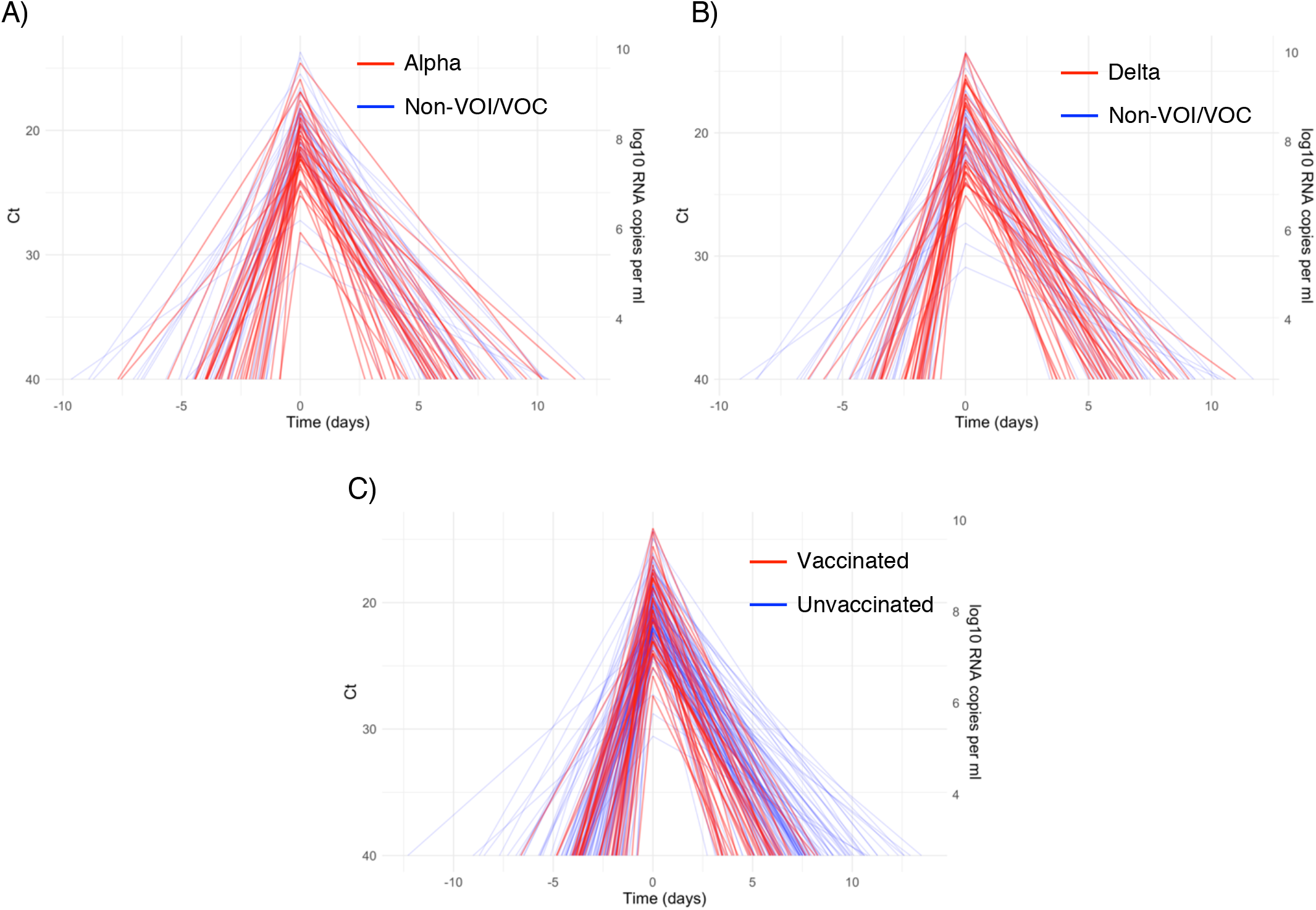
Mean posterior viral trajectories for each person. Pane (A) depicts alpha infections (red) against non-VOI/VOC infections (blue). Pane (B) depicts delta infections (red) against non-VOI/VOC infections (blue). Pane (C) depicts infections in vaccinated people (red) against unvaccinated people (blue). Trajectories are aligned temporally to have the same peak time.

**Supplementary Figure 13.**
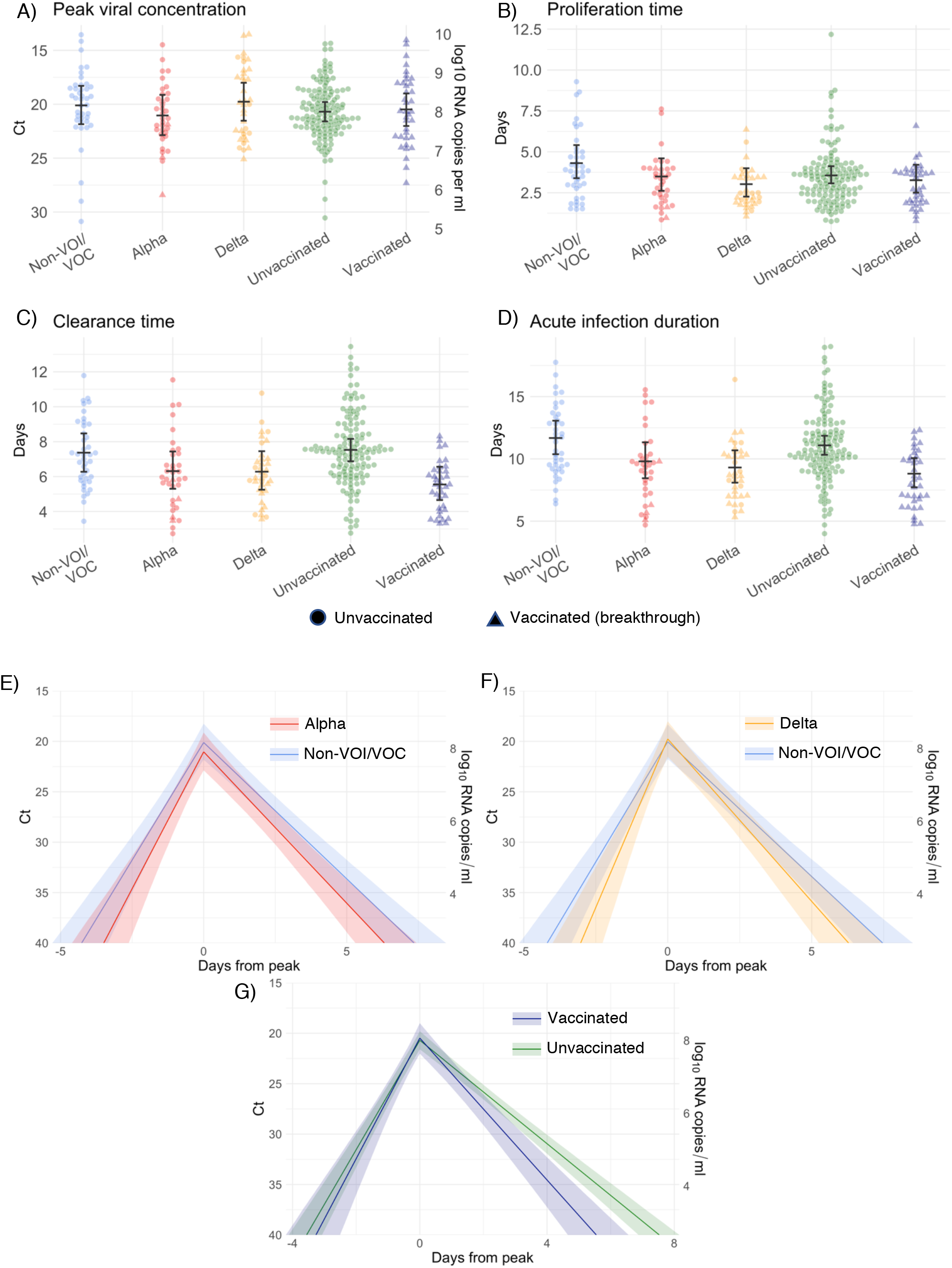
Estimated viral trajectory parameters for SARS-CoV-2 infections by variant and vaccination status using uninformative priors. Individual posterior means (points) with population means and 95% credible intervals (hatched lines) for (A) the peak viral concentration, (B) the proliferation duration, (C) the clearance duration, and (D) the total duration of acute infection for individuals infected with a non-VOI/VOC (blue), alpha (red), or delta (purple), and for individuals who were unvaccinated (green) or vaccinated (maroon). Circles denote unvaccinated individuals and triangles denote vaccinated individuals (breakthroughs). The points are jittered horizontally to avoid overlap. Solid lines in panes (E)-(F) depict the mean posterior viral trajectories for alpha (E, red) and delta (F, purple) infections respectively relative to non-VOI/VOC infections (blue), as specified by the population means and credible intervals in (A)-(D). Solid lines in pane (G) depict the mean posterior viral trajectory for vaccinated (maroon) relative to unvaccinated (green) individuals. The shaded regions in (E)-(G) represent 95% credible areas for the mean population trajectories. Priors were informed by a previous analysis and are defined in Eq. (S10).

**Supplementary Figure 14.**
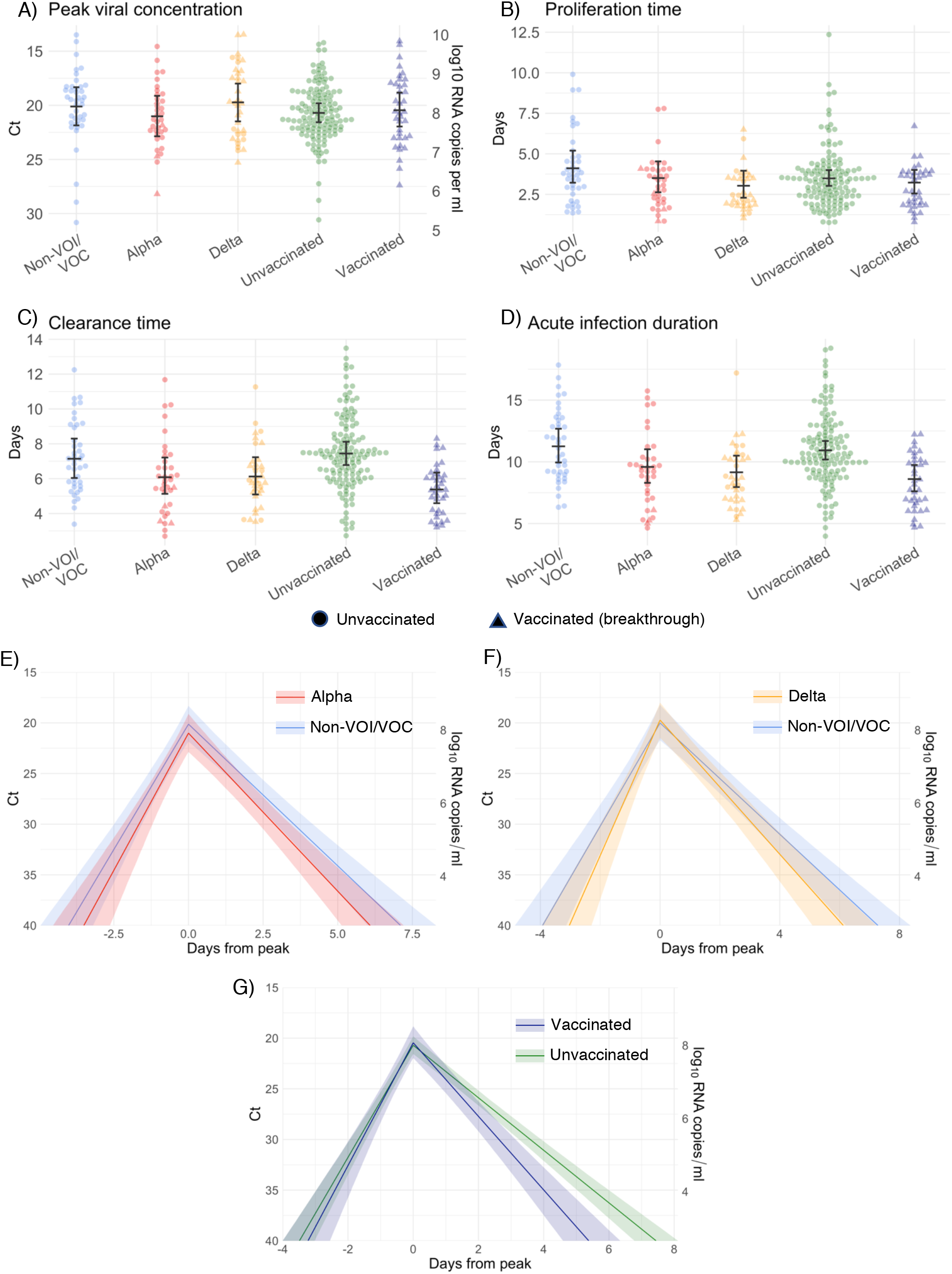
Estimated viral trajectory parameters for SARS-CoV-2 infections by variant and vaccination status using biased (low) priors. Individual posterior means (points) with population means and 95% credible intervals (hatched lines) for (A) the peak viral concentration, (B) the proliferation duration, (C) the clearance duration, and (D) the total duration of acute infection for individuals infected with a non-VOI/VOC (blue), alpha (red), or delta (purple), and for individuals who were unvaccinated (green) or vaccinated (maroon). Circles denote unvaccinated individuals and triangles denote vaccinated individuals (breakthroughs). The points are jittered horizontally to avoid overlap. Solid lines in panes (E)-(F) depict the mean posterior viral trajectories for alpha (E, red) and delta (F, purple) infections respectively relative to non-VOI/VOC infections (blue), as specified by the population means and credible intervals in (A)-(D). Solid lines in pane (G) depict the mean posterior viral trajectory for vaccinated (maroon) relative to unvaccinated (green) individuals. The shaded regions in (E)-(G) represent 95% credible areas for the mean population trajectories. Priors were chosen to be unrealistically low and are defined in Eq. (S11).

